# Deep learning-driven risk-based subtyping of cognitively impaired individuals

**DOI:** 10.1101/2021.12.08.21267495

**Authors:** Michael F. Romano, Xiao Zhou, Akshara R. Balachandra, Michalina F. Jadick, Shangran Qiu, Diya A. Nijhawan, Prajakta S. Joshi, Peter H. Lee, Maximilian J. Smith, Aaron B. Paul, Asim Z. Mian, Juan E. Small, Sang P. Chin, Rhoda Au, Vijaya B. Kolachalama

## Abstract

Quantifying heterogeneity in Alzheimer’s disease (AD) risk is critical for individualized care and management. Recent attempts to assess AD heterogeneity have used structural (magnetic resonance imaging (MRI)-based) or functional (Aβ or tau) imaging, which focused on generating quartets of atrophy patterns and protein spreading, respectively. Here we present a computational framework that facilitated the identification of subtypes based on their risk of progression to AD. We used cerebrospinal fluid (CSF) measures of Aβ from the Alzheimer’s Disease Neuroimaging Initiative (ADNI) (n=544, discovery cohort) as well as the National Alzheimer’s Coordinating Center (NACC) (n=508, validation cohort), and risk-stratified individuals with mild cognitive impairment (MCI) into quartiles (high-risk (H), intermediate-high risk (IH), intermediate-low risk (IL), and low-risk (L)). Patients were divided into subgroups utilizing patterns of brain atrophy found in each of these risk-stratified quartiles. We found H subjects to have a greater risk of AD progression compared to the other subtypes at 2- and 4-years in both the discovery and validation cohorts (ADNI: H subtype versus all others, p < 0.05 at 2 and 4 years; NACC: H vs. IL and LR at 2 years, p < 0.05, and a trend toward higher risk vs. IH, and p < 0.05 vs. IH, and L risk groups at 48 months with a trend toward lower survival vs. IL). Using MRI-based neural models that fused various deep neural networks with survival analysis, we then predicted MCI to AD conversion. We used these models to identify subtype-specific regions that demonstrate the largest levels of *atrophy-related importance*, which had minimal overlap (Average pairwise Jaccard Similarity in regions between the top 5 subtypes, 0.25±0.05 (± std)). Neuropathologic changes characteristic of AD were present across all subtypes in comparable proportions (Chi-square test, p>0.05 for differences in ADNC, n=31). Our risk-based approach to subtyping individuals provides an objective means to intervene and tailor care management strategies at early stages of cognitive decline.

## Introduction

The projected cost of caring for tens of millions of individuals who have Alzheimer’s disease (AD) worldwide is going to exceed trillions of dollars over the next few decades (Anonymous 2020). Financial estimates, however, cannot measure the physical, health, emotional and psychological strain of AD on family members and caregivers. There is a pressing need to identify persons at risk of progressing to AD from subclinical AD or mild cognitive impairment (MCI), as they may benefit the most from early interventions and management. Importantly, as disease-modifying therapies are undergoing regulatory scrutiny (Knopman et al. 2021; Robinson 2021), it becomes increasingly important to identify those who would benefit from such interventions.

Not all persons with MCI develop AD. To this end, several novel frameworks have been constructed to identify individuals with normal cognition or MCI who will progress to AD using both imaging (Ding et al. 2019; Iddi et al. 2019; Lin et al. 2018; McRae-McKee et al. 2019) and cerebrospinal fluid (CSF) biomarkers (Buchhave 2012; Fagan et al. 2007; Li et al. 2016; Lista et al. 2014; Michaud et al. 2015). Studies have also focused on subtyping AD at fixed points in time based on structural and functional imaging (ten Kate et al. 2018; Vogel et al. 2021). Finally, Young et al. (2018) introduced a technique that modeled AD across static subtypes and dynamically over time. While these studies have underscored the importance of characterizing AD heterogeneity, structure- or function-based subtyping does not always translate to precise identification of patients who manifest with AD. One needs to rather pursue this task using a bottom-up strategy wherein the subtyping is defined based on the disease outcome itself. This would enable better tracking of disease progression and quantifying an individual’s make-up at their symptomatic origins, followed by binning them as rapid or slow progressors as well as non-progressors. We contend that a risk-based strategy of subtyping AD using an instantaneously observed biomarker (i.e., CSF) would allow for the identification of patients who are likely to progress to AD. CSF levels of amyloid-β (Aβ) have been widely used to predict progression to AD (Anonymous; Blennow et al. 2010; Heister et al. 2011; Lista et al. 2014; Michaud et al. 2015; Visser et al. 2009). While these fluid-based measures have substantial inter-lab variability, standardized datasets, such as that of the Alzheimer’s Disease Neuroimaging Initiative (ADNI), maybe more reliable (Lista et al. 2014). CSF-based subtyping could enable precise targeting of patients who would benefit from novel therapeutic agents such as biologics, allowing physicians to intervene before their patients experience potentially irreversible changes in their cognition (Blennow et al. 2010; Heister et al. 2011; Lista et al. 2014; Michaud et al. 2015; Visser et al. 2009). By mapping these highly standardized CSF datasets to their corresponding MRIs, we could apply more robust deep learning techniques to predict risk in outside datasets, which would be challenging to do with CSF biomarkers alone.

We fused deep learning with classical survival analysis to integrate spatial information from MRIs and temporal information and estimate precise forecasts of progression from MCI to AD. Recently, survival methodologies have been integrated into deep learning, including Cox proportional hazard-type models (Katzman et al. 2018; Kim et al. 2019; Mobadersany et al. 2018; Wulczyn et al. 2021; Zhu et al. 2016) and more flexible variants such as Nnet-survival, which do not require the proportional hazards assumption (Gensheimer and Narasimhan 2019; Zhang 2020). We hypothesized that models that combine flexible survival prediction in conjunction with convolutional neural networks and minimally processed T1-weighted MRI that utilize region-specific gray matter volumes would be ideal for measuring the predictive value of each of these risk-based subtypes. Using this rich framework, we (1) performed subtyping of persons based on a well-identified and standard measure of progression to AD (i.e., CSF Aβ); (2) used this stratification to identify patterns of atrophy that characterize each subtype; (3) established the utility of this stratification scheme by training a machine-learning model on our data that forecasts progression from MCI to AD in a granular fashion; and finally (4) utilized neuropathology as a reference standard to confirm a diagnosis of AD.

## Methods

### Study population and data selection

Data were collected from the Alzheimer’s Disease Neuroimaging Initiative (ADNI) database for training, internal validation, and internal testing, and from the National Alzheimer’s Coordinating Center (NACC) for external validation.

### ADNI cohort

ADNI comprises a multi-center, longitudinal study with an overall goal of facilitating the development of novel therapeutics by identifying biomarkers that identify and portend progression of AD. Thus far, there have been 4 separate phases of ADNI. For this dataset, visits for all subjects were selected from the person registry with a last user date of April 9, 2020; this includes subjects enrolled in all different phases of ADNI, including ADNI 1, ADNI GO, ADNI2, and ADNI 3. General requirements for all phases included persons between 55-90 years old, a partner able to be present for collateral, a Geriatric Depression Scale less than 6, and fluency in one of Spanish or English. Mild cognitive impairment (MCI) was defined by ADNI similarly across all 4 phases. To qualify as MCI, consistent criteria included the following: a person had to have 1) a complaint about some aspect of cognition; 2) mini-mental state exam (MMSE) score ≥ 24 and a clinical dementia rating (CDR) equal to 0.5 with preserved daily function; and crucially, 3) some measured memory loss based on a Logical Memory test, adjusted for years of education. Persons had to have the amnestic domain affected to be enrolled. To meet criteria for AD, a person had to have a CDR ≥ 0.5, MMSE ≤ 26, an abnormal Logical Memory test, and meet criteria for AD based on NINCDS-ADRDA criteria for probable AD (McKhann et al. 1984).

### ADNI pre-processing

To group together visits for persons from different data files, we merged visit data using as merge codes the person ID (‘RID’), and the visit codes (‘VISCODE’ and ‘VISCODE2’, where available). Visits labeled as ‘f’, ‘nv’, ‘uns1’, and ‘tau’ were excluded from consideration. Diagnoses were established for persons using ‘DXCURREN’, ‘DXCHANGE’, ‘‘DXCHANGE’, and ‘DIAGNOSIS’ fields from the diagnosis summary file, corresponding to phases of the study ‘ADNI1’, ‘ADNIGO’, ‘ADNI2’, and ‘ADNI3’, while confirmation of a diagnosis of dementia due to AD was confirmed with the ‘DXAD’ field for ADNI1, and the ‘DXDDUE’ field for the remainder of the phases. Data from baseline and screening visits were combined, taking the baseline diagnosis result and other baseline assessments first, and filling in the remainder of the data with assessments conducted exclusively at a screening visit. From the collected data, visits where persons had a 3 Tesla T1-weighted MRI scan (between 2.7 and 3.1 Tesla), CSF data collected, and a diagnosis of mild cognitive impairment (either late or early mild cognitive impairment where specified), as made by clinicians using multimodal criteria specified by ADNI, were identified. The first such visit was identified for each person.

### ADNI image selection

Raw MRI images on the ADNI database were queried using the keyword arguments *MP*RAGE* and *SPGR*, corresponding to magnetization-prepared rapid gradient-echo and spoiled gradient-recalled echo, respectively. Magnet strengths between 2.9 and 3.1 were queried. Once downloaded, images were first filtered by the desired date for each person’s MCI visit. Then, the Mayo Clinic quality control information was used to further inform which image to use. If there was more than a single image at a given visit for each person, images were selectively kept using the following criteria in the following order of importance: being fully sampled (i.e. the image description did not contain the phrases “SENSE”, “ACCEL”, or “GRAPPA”); receiving a “pass” on the Mayo Clinic quality control sheet where this information was available; being taken at the most recent date; being chosen in the Mayo Clinic quality control sheet as “selected”; and finally, if there was still more than a single scan remaining, the image with the highest image ID, which generally corresponded to the latest image obtained in a series, was taken. If at any point application of these criteria led to removal of all scans for a given subject, the step was skipped to keep as many scans as possible (for example, if a subject only had accelerated MRI scans, accelerated MRI scans were used for that subject). Overall, 49 persons were selected from ADNI1 (45 at the baseline visit, 1 each at month 60, 96, 108, and 120), 113 from ADNIGO (112 at the baseline visit, 1 at month 24), 321 from ADNI2 (309 at baseline visit, 11 at month 24, and 1 at month 48), and 59 from ADNI3 (all at baseline visit), yielding 544 total participants (as described later, 1 subject was removed from analysis due to poor image co-registration). Following an analysis of patient demographics and clustering of patients into different anatomical subtypes (Figures 1 and 2), 540 participants were randomly selected for the remainder of the study for the number of participants to be evenly divided when conducting the 5-fold cross validation for our deep-learning models. This image selection is illustrated in Supplementary Figure 1.

**Figure 1.**
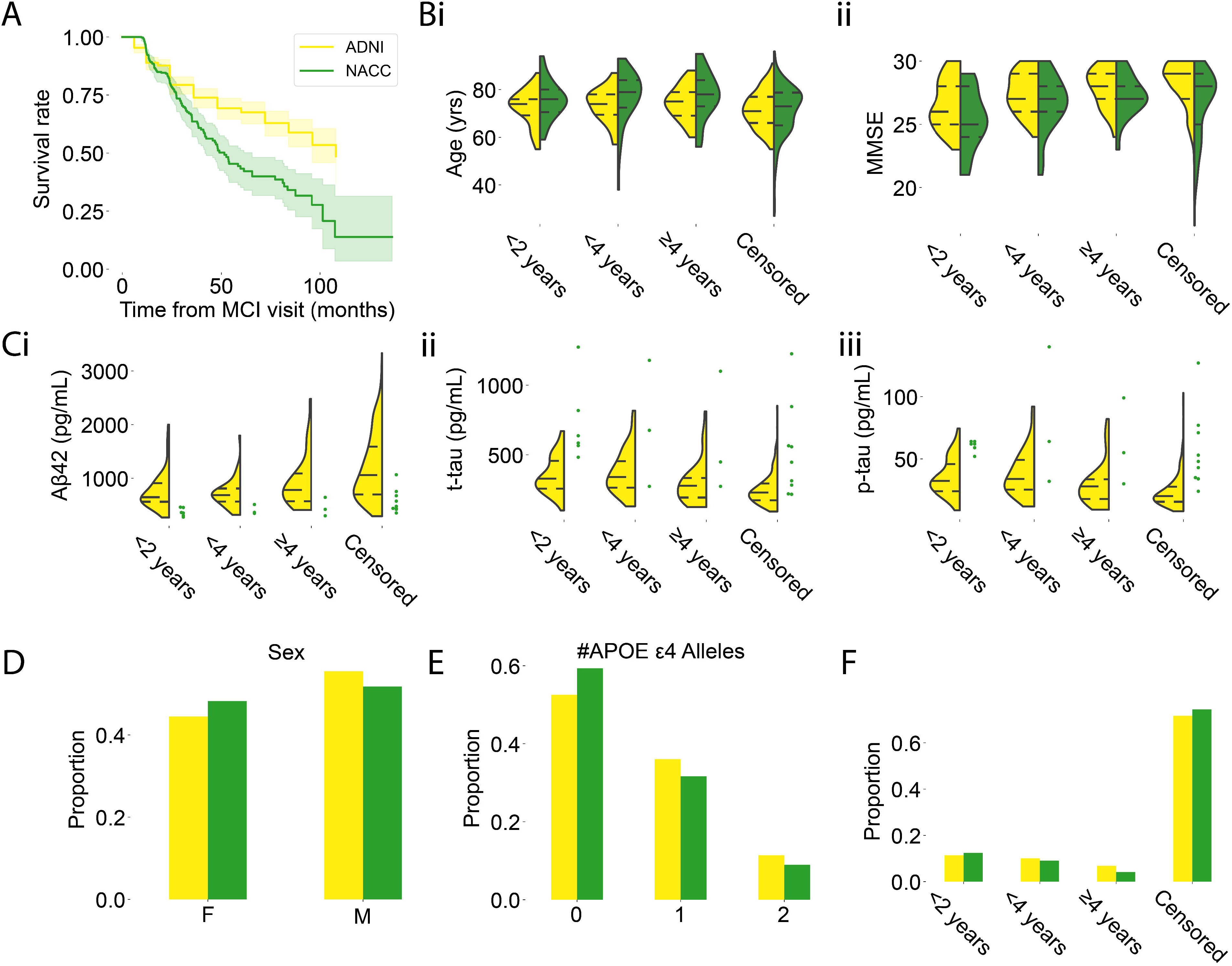
Study population. Summary statistics of clinical and demographic parameters of persons from the Alzheimer’s disease neuroimaging initiative (ADNI) and National Alzheimer’s coordinating center (NACC) cohorts are shown. (A) Kaplan-Meier survival curves with 95% confidence intervals computed for our two populations (ADNI: n=544, 390 right-censored; NACC: n=508, 378 right-censored). Survival (time to progression from mild cognitive impairment to Alzheimer’s disease) was approximately the same for both cohorts at 24 months (X^2^(1)=0.06, p=0.80) though survival in the ADNI group exceeded that of the NACC group at 48 months (X^2^(1)=14.56, p<0.005, n= 544 total observations, 390 right-censored observations for ADNI, n= 508 total observations, 378 right-censored observations for NACC). **B.** Differences in age (i) and mini-mental status exam score (ii) for persons in the NACC and ADNI datasets. MMSE scores in the ≥ 4 year and censored groups exceeded that in the < 2 years groups in the ADNI dataset, while MMSEs in the censored group exceeded both <2 year and < 4 year groups (Kruskall-Wallis test, H=39.8, df=3, p=1.1e-08; Dunn test post-hoc with Bonferroni correction: < 2 years vs censored, p < 0.01; < 4 years vs censored, p=0.016; <2 years vs ≥ 4 years, 0.018). This suggests a general pattern of higher MMSE scores corresponding with longer time-to-progression. In addition, censored persons demonstrated similar MMSE scores as the ≥ 4 year group (p=1). The NACC cohort demonstrated a similar trend, with MMSE scores in the censored group exceeding those in the < 2 year group (Kruskal-Wallis test, H=9.93, df=3, p=0.019; n_<2 years_=112, n_<4 years_=34, n_≥4 years_=21, n_Censored_=33, n_missing_=266, 12, 0, and 30, respectively; Dunn post-hoc with Bonferroni-correction, p=0.017) and MMSE scores in the ≥4 year group approaching significance relative to the <2 year group (p=0.074). Overall, the NACC cohort demonstrated lower MMSE scores than the ADNI cohort (w=72597.5, p=1.2e-12, n_ADNI_=544, n_NACC_=200, n_missing,NAC_=308, n_missing,ADNI_=0). Persons in the ADNI group were also, on average, younger than the NACC group (Wilcoxon rank-sum test, w=127436, p=0.029, n_NACC_=508, n_ADNI_=544). In both datasets, the age of persons in the censored group trended toward being the youngest out of all groups, though no groups demonstrated differences when controlled for multiple comparisons in ADNI (Kruskal-Wallis test, H=9.06, df=3, p=0.028, n_<2 years_=390, n_<4 years_=55, n_≥4 years_=37, n_Censored_=62) and only the comparison between the <4 year group and the control group demonstrated a significant difference in the NACC cohort (Kruskal-Wallis test, H=25.8, df=3, p=1.05e-05, n_<2 years_=378, n_<4 years_=46, n_≥4 years_=21, n_Censored_=63; <4 years vs Censored: p=0.00026). **C.** Concentration profiles of three different CSF biomarkers in the two cohorts. Statistics not computed for the NACC dataset due to the large amount of missing data. For t-tau, persons in the ≥ 4 year group and censored group demonstrated larger concentrations (Kruskal-Wallis test, H=71.0, df=3, p=2.65e-15, n_<2 years_=390, n_<4 years_=55, n_≥4 years_=37, n_Censored_=62; ≥ 4 years vs < 2 years, p=0.043, ≥4 years vs <4 years, 0.026, censored vs <2 years, p=1.71e-09, censored vs <4 years, p=1.49e-09). A similar pattern was observed for p-tau (Kruskal-Wallis test, H=79.4, df=3, p=4.06e-17, n_<2 years_=390, n_<4 years_=55, n_≥4 years_=37, n_Censored_=62; ≥ 4 years vs < 2 years, p=0.042, ≥4 years vs <4 years, 0.023, censored vs <2 years, p=1.73e-10, censored vs <4 years, p=1.29e-10). For Aβ we observed similar CSF concentrations for <2 year, <4 year, and ≥4 year groups, while the censored groups demonstrated relatively higher levels (Kruskal-Wallis test, H=61.8, df=3, p=2.4e-13, n_<2 years_=390, n_<4 years_=55, n_≥4 years_=37, n_Censored_=62; censored vs <2 years, p=4.4e-08, censored vs <4 years, p=3.5e-08). **D,E,F.** There were no differences in distributions of sex (X^2^(3)=1.33, p=0.25), APOe4 status (X^2^(3)=4.37, p=0.11), or proportion of persons in each of the progression groups between the two datasets (X^2^(3)=4.18, p=0.24).

**Figure 2.**
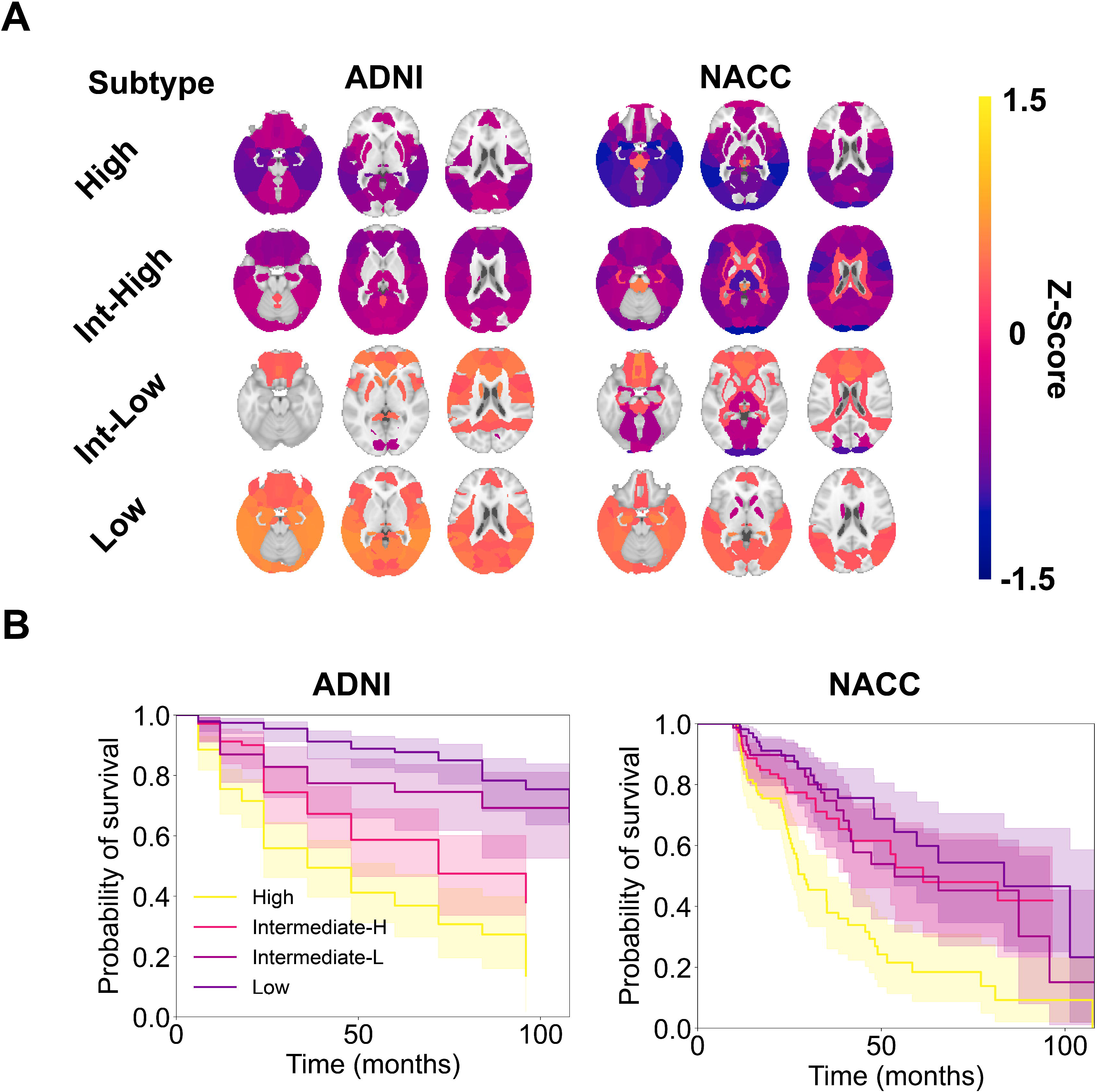
Patterns of risk-based subtypes during MCI to AD conversion. **A.** Mean z-scored gray matter in different brain regions for high (H), intermediate-high (I-H), intermediate-low (I-L), and low (L) risk subtypes (n=120 patients with both MCI and AD visits). How “hot” each region is signifying the z-scored, normalized gray matter volume for that subregion over all subjects in that subtype. The mean z-scored gray matter volumes show high correlation within-subtypes over time, and less correlated with other subtypes (H vs H, ρ=0.94, p=2.4e-31; I-H vs I-H, ρ=0.73, p=2.5e-12; I-L vs I-L, ρ=0.74, p=1.7e-12; L vs L, ρ=0.61, p=4.1e-08). **B.** A Sankey plot demonstrating flow from subtypes at each person’s MCI visit (left) to subtypes at each person’s AD visit (right). The plurality of persons from each subtype transitioned to the same subtype at their AD visit based on gray matter volume (59/60 high risk persons, 19/29 intermediate-high risk persons, 9/13 intermediate-low risk persons, and 9/18 low risk persons). **C.** When grouped by lobe assignment, subtypes demonstrate similar levels of normalized gray-matter atrophy in characteristic regions at both the MCI and AD time points (n_ADNI_=540, n_ADNI, AD_=120).

### ADNI image curation for AD-visits

An analogous process was used for Alzheimer’s Dementia visits (Fig. 3). Visits were selected where during the first visit where a patient was diagnosed with Alzheimer’s Dementia, an MRI at this date was also available. At this point, a similar image selection criterion was used as described in *ADNI image selection*. In sum, 123 patients in the ADNI cohort had an AD visit that corresponded to their MCI visit, though only 120 patients had scans that were not corrupted during download and were used for further analysis.

**Figure 3.**
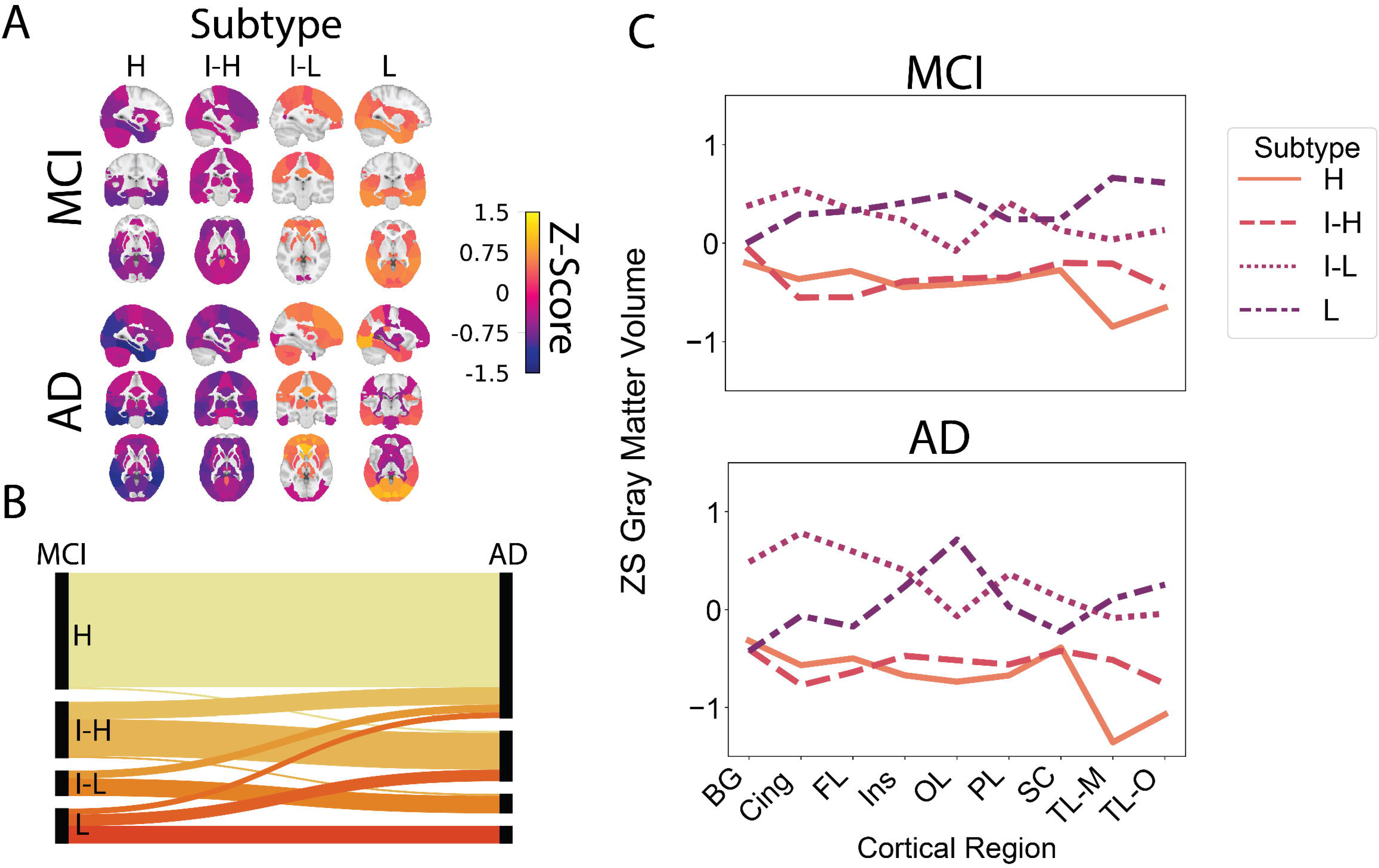
Distribution of risk-based subtypes. **A**. Gray matter volumes for persons in each sub-group are highly correlated across subtypes (Spearman’s correlation coefficient, high-risk subtype in ADNI vs high-risk subtype in NACC, ρ=0.85, p=7.5e-20; intermediate-high risk in ADNI vs intermediate-high risk in NACC, ρ=0.73, p=2.5e-12; intermediate-low risk in ADNI vs intermediate-low risk in NACC, ρ=0.70, p=8e-11; ADNI low-risk vs NACC low-risk, ρ=0.87, p=2.7e-21). Hotter colors indicate larger z-scored, normalized gray matter volumes, z-scored to the mean and standard deviations of each region in the complete ADNI dataset (n=544). Z-scores were thresholded at a value of 0.3. **B.** Survival curves for persons in each of the subtypes in the ADNI and NACC dataset, compared at time points 24, 48, and 96 months. In the ADNI dataset, the high-risk subtype demonstrates a higher risk of progression to AD than the intermediate-high risk subtype at times t=24 months and t=48 months (X^2^(1)=7.02, p=0.008, X^2^(1)=4.84, p=0.028 (Benjamini-Hochberg corrected)). Intermediate-high risk and intermediate-low risk subtypes do not differ in risk of progression at t=24 months (X^2^(1)=1.71, p=0.19) but do differ at 48 months (X^2^(1)=5.59, p=0.018). Intermediate-low and low-risk subtypes differ at both 24 and 48 months (X^2^(1)=10.0, p=0.0016; X^2^(1)=4.81, p=0.028). In the NACC dataset, the high-risk subtype demonstrates a higher risk of progression to AD at time t=24 months than both intermediate-low risk and low risk subtypes (X^2^(1)=7.50, p=0.018, X^2^(1)=9.85, p=0.010), and a trend to higher risk of progression to AD than the intermediate-high risk subtype (X^2^(1)=2.96, p=0.13). At time t=48 months, the high-risk subtype demonstrates a higher risk of progression than the intermediate-high risk and low risk subtypes (X^2^(1)=9.49, p=0.0062; X^2^(1)=13.8, p=0.00020), and a strong trend compared with the intermediate-low risk subtype (X^2^(1)=4.71, p=0.060) Intermediate-high risk and intermediate-low risk subtypes do not differ in risk of progression at t=24 months (X^2^(1)=1.71, p=0.19) but do differ at 48 months (X^2^(1)=5.59, p=0.18). Intermediate-low and low-risk subtypes differ at both 24 and 48 months (X^2^(1)=10.0, p=0.0016; X^2^(1)=4.81, p=0.028). Intermediate-high, intermediate-low, and low risk subtypes did not demonstrate differences from one another at any time points.

### ADNI image curation for pre-training

Finally, for pre-training our deep-learning models, we constructed a dataset consisting of MRIs from unused patients in the ADNI cohort. These consisted of all 3-Tesla MRI images, IR-(F)SPGR and MP-RAGE, that we obtained at our collection date in the ADNI dataset, for patients that were *not* used for the main part of the study. Images that did not have diagnoses at the time of the visit were excluded. Image curation and metadata pre-processing for ADNI is shown in Supplementary Figure 1 for reference.

### NACC cohort

The NACC database hosts a Uniform Data Set (UDS) comprised of longitudinal data, collected from persons in National Institute on Aging Alzheimer’s Disease Research Centers (ADRCs), each with its own protocol for enrollment, and each with its own protocol for diagnosis of disease (a team of physicians versus a single physician). Diagnoses for MCI and AD are primarily made based on clinician judgement. MCI persons are defined as those with preserved day to day function, though with a concern from the person, person’s partner, or physician about the person’s cognition and impairment in at least one cognitive domain. Dementia is diagnosed by a measured and clinically determined progressive decline in cognitive ability with impacted day-to-day function, in addition to impairment in at least one of 5 cognitive domains. AD is determined by clinical judgment based on available data.

Subjects in our study were selected from a data freeze on December 12, 2020. For each subject, visits where the subject had mild cognitive impairment (amnestic or non-amnestic, single or multiple domain) were identified. Out of all visits for each patient who carried a diagnosis of mild cognitive impairment, the visit that was closest to a date at which they had a 3T, T1-weighted MRI was kept. If the time between the clinical visit and MRI was longer than 6 months, the patient was dropped from consideration. CSF values were assigned to the nearest diagnostic visit provided the visit occurred within ±6 months. CSF values in the NACC dataset were all obtained via an ELISA assay method (total of 21 samples).

Metadata for each of the T1-weighted scans were used to select which T1-weighted MRI to use out of the several MRIs available for each visit. Only three-dimensional, original, SPGR or MP RAGE images were used. Their single smallest dimension had to be at least 80 voxels. If a person had fully sampled scans, these were selected in place of any accelerated scans such as GRAPPA or SENSE. Finally, if there was more than a single image left for a person, an image collected that met the criteria was selected at random but with preference to the most recently acquired scans. Image curation and metadata pre-processing for NACC is shown in Supplementary Figure 2 for reference.

### Image registration, normalization, and segmentation

Images were parcellated utilizing SPM 12 software version 7771, CAT12.7 version 1728 and MATLAB version 2020a Update 4, and an overall outline of the processes are demonstrated in Supplementary Figure 3. Images were batch processed using the following methodology. First, MRIs were centered such that the center of each scan was located at the image origin. This was accomplished using a script generously provided by Dr. Landau and Alice Murphy of the Helen Wills Neuroscience Institute at University of California, Berkeley.

At this point, images were further processed in one of two ways, depending on the deep learning pipeline that was to follow.

For the multi-layer perceptron model, a parcellation pipeline that utilized CAT12.7 (Computational Anatomy Toolbox, http://www.neuro.uni-jena.de/cat/) was used in order to determine normalized gray matter (GM) volumes for pre-specified regions of the brain. These normalized GM volumes were then used as input for the multi-layer perceptron model. In order to obtain these normalized gray matter volumes, recentered MRIs were first batch processed using CAT12 segmentation with the default parameters. The Neuromorphometrics atlas was used for volumetric analysis (from MICCAI 2012 Grand Challenge and Workshop on Multi-Atlas Labeling, using MRIs from the OASIS project, labeled data provided by Neuromorphometrics, Inc. (http://Neuromorphometrics.com/) under academic subscription); the original atlas was constructed from brains originally obtained from OASIS (https://www.oasis-brains.org/). This resulted in 544 gray matter volumes for the ADNI dataset and 508 gray matter volumes for the NACC dataset. Gray matter volumes were averaged across hemispheres. These volumes were normalized by dividing by the total intracranial volume, yielding normalized gray matter volumes. Normalized gray matter volumes are referred to simply as gray matter volumes throughout the main text of the manuscript. The regions corresponding to ventricles were removed for further analysis (CSF, 3rd ventricle, 4th ventricle, inferior lateral ventricle, and lateral ventricle).

For other models, we used a standard SPM12 (Statistical Parametric Mapping, https://www.fil.ion.ucl.ac.uk/spm/software/spm12/) pipeline which consisted of co-registering and masking each brain using the following steps: 1. Segmentation of each brain into gray matter, white matter, and CSF; 2. Bias-correction of each brain; 3. Normalization of each bias-corrected brain into MNI (Montreal Neurological Institute) space using the deformation field obtained from (1); 4. Masking each brain by thresholding the sum of the gray matter, white matter, and CSF probability atlases at a value of 0.2, and taking the pointwise product of the normalized brain and the thresholded, normalized atlas. The outputs were saved as 64-bit floating point numbers in Nifti files. During the quality control process, we found the brain of 1 subject to be poorly registered. This subject was removed from all analyses.

### Empirical survival analyses and demographics plotting

For survival analyses, the *lifelines* package was relied upon extensively (Davidson-Pilon 2019). The class KaplanMeierFitter was used to plot empirical Kaplan-Meier plots, CoxPHFitter was used to compute proportional hazard tests, and the function *survival_differences_at_fixed_point_in_time_test* was used to compute differences in survival at different time points. *Seaborn* was used extensively for scatter plots and violin plots as well (Waskom 2021), and *Scipy* was used to compute Kruskal-Wallis tests, Chi-square tests, and Mann-Whitney U tests (Virtanen et al. 2020). Bar plots were created using *pandas* version 1.0.1.

### Subtype analysis

#### Centroid computation

Subtyping patients was performed as follows. As has been argued by Buchhave (2012), levels of Aβ-42 have been shown to change up to 10 years prior to a diagnosis of Alzheimer’s Disease. We therefore divided CSF values for all cases in the ADNI cohort by Aβ quartile to risk-stratify each case into high-risk, intermediate-high risk, intermediate-low risk, and low-risk categories corresponding to the lowest concentration of CSF Aβ through the highest concentration of CSF Aβ, respectively. The gray matter volumes for each region for each participant in each of these subtypes were then obtained. For each region across all subtypes, the mean and standard deviation of the gray matter volume in that region was computed. Gray matter volumes for each brain region were Z-scored to the mean and standard deviations of that respective brain region. In order to obtain centroids for each subtype, Z-scored gray matter volumes were averaged across all brains in the ADNI cohort preliminarily sorted into each of the 4 subtypes, for a total of 4 centroids. Centroids were computed using the entire 544 patient ADNI dataset.

### Subtype assignment

In order to assign final subtypes, ADNI and NACC brain regions were all Z-scored using the respective region means and region standard deviations obtained as specified above. Spearman’s correlation coefficient was then calculated between each brain and each of the 4 subtype centroids. The subtype corresponding to the highest correlation coefficient was assigned as the subtype for that patient. Subtypes were assigned in the same way for the images obtained at the AD visit. To plot the Sankey Diagram demonstrating changes in subtype over time (Figure 2B), we utilized the python library *plotly* (https://plotly.com/). Brains were plotted using Nilearn (https://nilearn.github.io).

In order to compute statistics to compare differences in normalized gray matter volumes between different regions and between different subtypes, we utilized a linear mixed-effects model in the package lme4 version 1.1-27.1 in R, and performed post-hoc analysis using lmerTest version 3.1-3 in R. The model was constructed as:

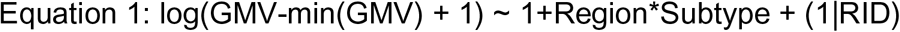

where GMV is normalized gray matter volume, RID is the individual subject identifier, and Region and Subtype are self-explanatory. A log transformation was taken to reduce the skewness of the data, and a constant shift of the data was applied to make the data positive before taking a log transform. Diagnostics can be seen in Supplementary Figure 6. We utilized the least squares mean estimate for each brain region provided by *lsmeans* in lmerTest to rank GMV. A Dunn-Sidak correction was applied to examine pairwise comparisons between subtypes both in terms of overall gray matter volume and in terms of region-specific gray matter volume.

### Assignment of parcellated regions to lobe

Regions were assigned to lobes as designated in the CAT12 Neuromorphometrics Atlas assignments, with several exceptions. Specifically, regions in the frontal, parietal, and occipital lobe were each grouped together. The temporal lobe was split into mesial and non-mesial temporal lobe, where the mesial temporal lobe consisted of the entorhinal area, parahippocampal gyrus, hippocampus, and amygdala. The remainder of the area designated as “subcortical” was divided into subcortical and regions in the basal ganglia. Another linear mixed-effects model was constructed here for analysis as follows:

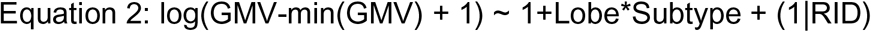

### Deep learning framework

#### Multi-layer perceptron

A multi-layer perceptron was utilized to predict a person’s risk of AD progression over time. The data was split in a 3:1:1 fashion (training:validation:testing), and 5-fold cross validation was used in order to ensure that each subject was used as test data exactly once, validation data exactly once, and training data three times. Normalized gray matter volume was utilized as input for this model. The model was repeated after adding age, MMSE score, and age together with MMSE score. Finally, a model was constructed with the only input being the three CSF biomarkers.

The model architecture consisted of a batch-normalization layer, followed by dropout, batch-normalization and leaky rectified linear-unit layers. This output was fed into a final linear layer and then to a sigmoid layer, which computed the *marginal probabilities of survival* in each of the three time bins: 0-24 months following the MCI visit, 24-48 months following the MCI visit, and 48-108 months following the MCI visit, all left-side inclusive. The model was saved when it had a lower total survival loss on the validation set. Please see *Survival Loss* in the appendix. Then, the model was tested on the external dataset, NACC. As the concordance index was highest for the simplest model (only parcellated brain regions) and the Brier score was comparable to the other models, this simpler model was used for testing.

To evaluate the success of the model, two statistics were computed: the concordance index (concordance_index_censored) and the Brier score (integrated_brier_score). The concordance index compares pairs of subjects and computes the proportion of pairs where our prediction of survival (i.e., which of the subjects “out-survives” the other) matches ground truth. To compute the concordance index, we calculated the predicted probability of survival at 24 months and used this to compare the predicted versus actual survival of two subjects. The Brier score is a statistic that measures the difference in survival for an individual against their predicted survival, and so measures bias of our predictions. These statistics were computed using scikit-survival (Pölsterl 2020). Using the marginal probabilities of survival, we can compute the overall probabilities of survival by taking the product of the marginal probabilities of surviving each of these time bins. Survival was interpolated for plotting using quadratic spline interpolation (Scipy).

#### Survival convolutional neural network

A modified convolutional neural network with survival loss function was trained to predict the risk of progression for patients in a time-based manner. As a result of the different loss function, it is referred to as the survival convolutional neural network (SCNN) in our work. In this model, 3D convolution is used to handle the volumetric MRI scans of size (121, 145, 121). The network contains 5 convolutional layers, each layer’s kernel size is set to be 3, with a stride of 1 and no padding. Batch normalization is applied throughout the 5 layers to prevent instability during training, and to decrease the number of epochs needed to achieve optimal state. After the normalization step, a leaky rectified linear unit is utilized as the activation function. Following this, a max-pooling layer of size 2 is attached to retrieve the most important features from previous output. In addition to L2 normalization (weight=0.01), dropout layers (probability=0.3) are also used to boost the robustness of the network. Finally, we flatten the output and apply fully connected layers for the final prediction of risks. Before training, we initialize the weights using the default initializer (Kaiming Uniform method). During the training, we feed the model with a batch size of 10 and use the stochastic gradient descent (SGD) optimizer with a learning rate of 0.01. Additionally, we calculated weights for each training sample based on their class frequency. The structure is visualized in Supplementary Figure 4.

Similar to the previous settings for experiments, the same datasets were used, as well as the (3:1:1) train:valid:test split (on ADNI dataset). NACC is again used as the external testing dataset. During training, the model is saved whenever it has a better concordance index on the validation set. Besides the concordance index, we also calculated the Brier score on all datasets for evaluation purposes. These two scores give assessments of accuracy (concordance index) in addition to calibration (Brier score). Multiple experiments with different parameters are made to achieve the best results; some of the most representative experiments are provided in the table.

#### Survival vision transformer

Transformers have been a hot topic recently and have been applied to various areas on different tasks. The vision transformer, for example, is one such application that is designed primarily for image-based tasks. In our work, we explored the vision transformer’s performance on 3D image handling in predicting the AD progression risk. Similar to the structure in (Dosovitskiy et al. 2020), we first split the 3D scan into smaller volumes of size (64, 64, 64), resulting in a number of patches, which is then sent into the mapper (linear mapper or convolutional mapper) to map into vectors with a dimension of 200. These vectors are then concatenated with a learnable class variable and encoded with relative locations using a positional encoder. The transformer will then take these vectors as input. The layers of the transformer are similar to the original structure: the input vectors will first be normalized and then used by multiple self-attention layers to compute the weighted sum, where the weights for each of them are based on the pairwise similarity between each element of the inputs; after that, the values will come with a skip-connection (which adds the original value to the computed values) and will be normalized again before being sent into the feed-forward layers. The outputs of these layers will come with skip-connection again. After the transformer, we append a multi-layer perceptron unit as the decoder to perform the risk prediction task. The structure is presented in Supplementary Figure 5.

We used the same data for training, validation, and testing -- same split, same bins, and same seeds for establishing a fair comparison. An SGD optimizer was used again when updating the weights, though with a smaller learning rate of 0.005. The dropout was set to 0.25, and the batch size was 5 (other sizes did not influence the results significantly). The weights were initialized using the Kaiming Uniform method as in the SCNN. We listed the 2 most representative results for SViT. In general, we observed that SViT performed worse than the SCNN.

#### SHAP Value Computation

SHaply Additive exPlanations (SHAP) values were utilized in order to determine the contribution of each input feature (normalized gray matter volume) to the predicted survival of each patient. SHAP values have been widely utilized (Lundberg et al. 2018; Verburg et al. 2021) in order to provide a measure of inference to machine-learning models. In our case, the DeepLIFT algorithm was used in order to compute a linear approximation whereby SHAP values are computed as the product of the backpropagated gradient for each feature, multiplied by the difference between the initial feature value and its expected value. For each fold, training data was used as the *background* data in order to compute the expected values for each feature. We used the *deepexplainer* method from the SHAP package to compute our values (https://github.com/slundberg/shap)).

#### Atrophy-Related Importance

SHAP values were sorted based on their Spearman correlation coefficients with Z-scored gray matter volumes. The regions with the 5 highest correlation coefficients for each subtype were selected for further analysis and plotted in Figure 5. We term these regions as having the largest *atrophy-related importance*, as their importance varies most directly with their normalized gray matter volume. To assess differences in the average SHAP values for these top regions within each subtype, we used a linear mixed effects model (lme4 package in R). Our model was specified as:

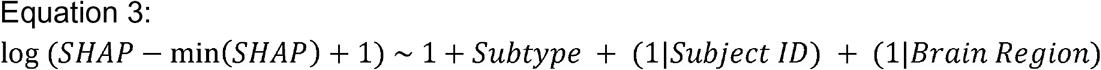

**Figure 4.**
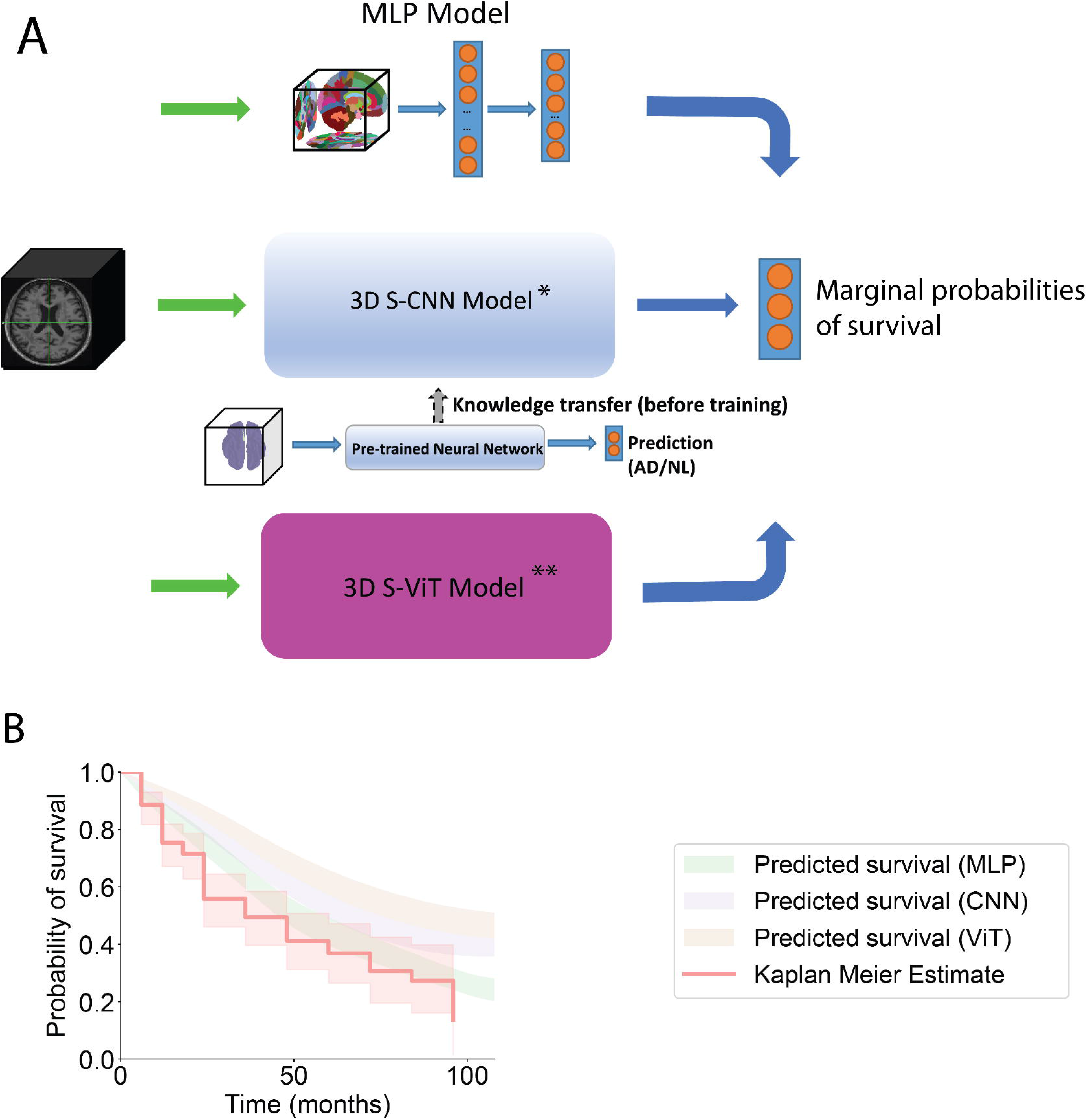
Schematics of deep learning frameworks. **A.** A schematic of a multilayer perceptron (MLP). Images were normalized to MNI space and segmented using CAT12, and gray matter volumes were obtained and normalized to the total intracranial volume for each subject. These values were fed into a multilayer perceptron with two fully connected layers and used to predict the marginal probability of survival up to 24, 48, and 108 months. A survival-convolutional neural network (S-CNN) was also constructed, with details in Supplementary Figure 4. This took as input the full, skull-stripped and normalized MRIs and was used to predict a similar output. Finally, a survival-vision transformer was constructed for comparison, with the same input and outputs as the S-CNN. **B.** An exemplar comparison of empirical survival curves (Kaplan-Meier estimate) and predicted survival curves (interpolated in 1-month increments using the marginal probabilities of survival from each model) for the high-risk subtype in the ADNI cohort. The MLP recapitulates the empirical survival curve best of all three models, as seen in Table 1.

**Figure 5.**
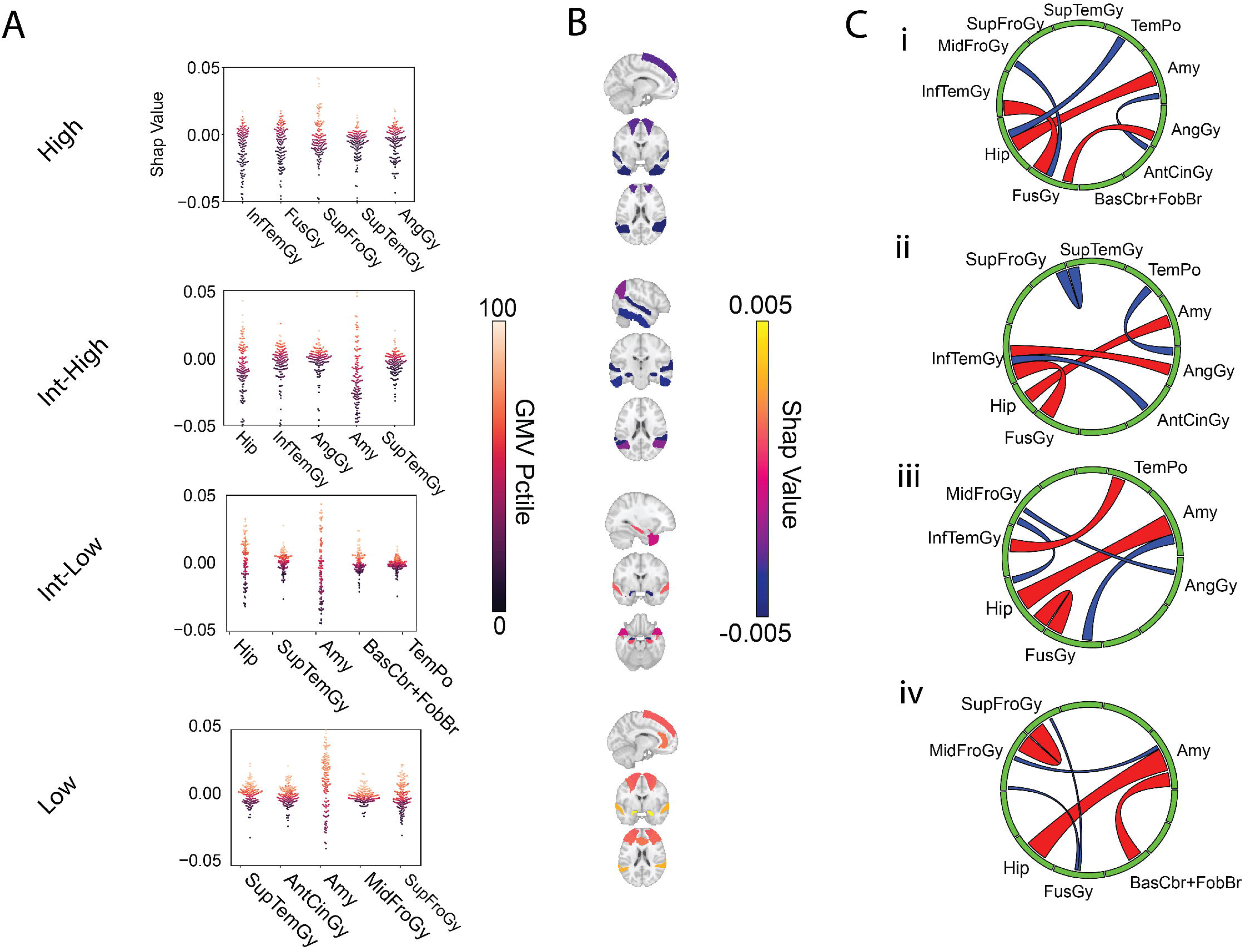
Spatial signatures of risk-based subtypes. Mean SHAP values from the multi-layer perceptron model across all 3 survival bins for the NACC dataset, and chord diagrams demonstrating large partial correlation coefficients between different regions. **A.** The 5 regions with the largest atrophy-modulated importance are plotted here for each subtype. Larger SHAP values correspond to more importance for that brain region in terms of predicting the average marginal probability of survival across the three time bins (0-24 months, 24-48 months, and ≥ 48 months), and hotter values correspond to larger gray matter volume (broken down into percentiles). SHAP values are on average, larger, for the top 5 regions in the low-risk subtype, which correspond to higher gray matter volumes and lower marginal probabilities of progression (i.e., higher probabilities of survival). SHAP values are, at the same time, smaller, for the top 5 regions in the high-risk subtype, corresponding to more largely negative SHAP values and correspondingly lower probabilities of progression. (Mixed-effects model, main effect of high-risk subtype vs intermediate-high risk subtype, z=-1.034, p=0.88; high risk subtype vs intermediate-low risk subtype, z=-3.008, p=0.0157; high risk vs low-risk subtype, p<0.001, Dunn-Sidak corrected for multiple comparisons). **B.** Brain masks with heat map overlays corresponding to average SHAP values across all individuals from each subtype for each of the top five regions highlighted in (A). **C.** Chord diagrams demonstrating subtype-specific signatures. Each edge represents a partial correlation coefficient between SHAP values between regions represented in (A). The 3 largest and 3 smallest partial correlations are displayed for each subtype. Red edges correspond to positive relationships, and blue edges correspond to negative relationships. Edge width corresponds to the magnitude of the partial correlation coefficient. Strong positive partial correlations between hippocampus and amygdala SHAP values are seen in all chord diagrams. Aside from this relationship, the remainder of the top relationships remain largely distinct.

**Figure 6.**
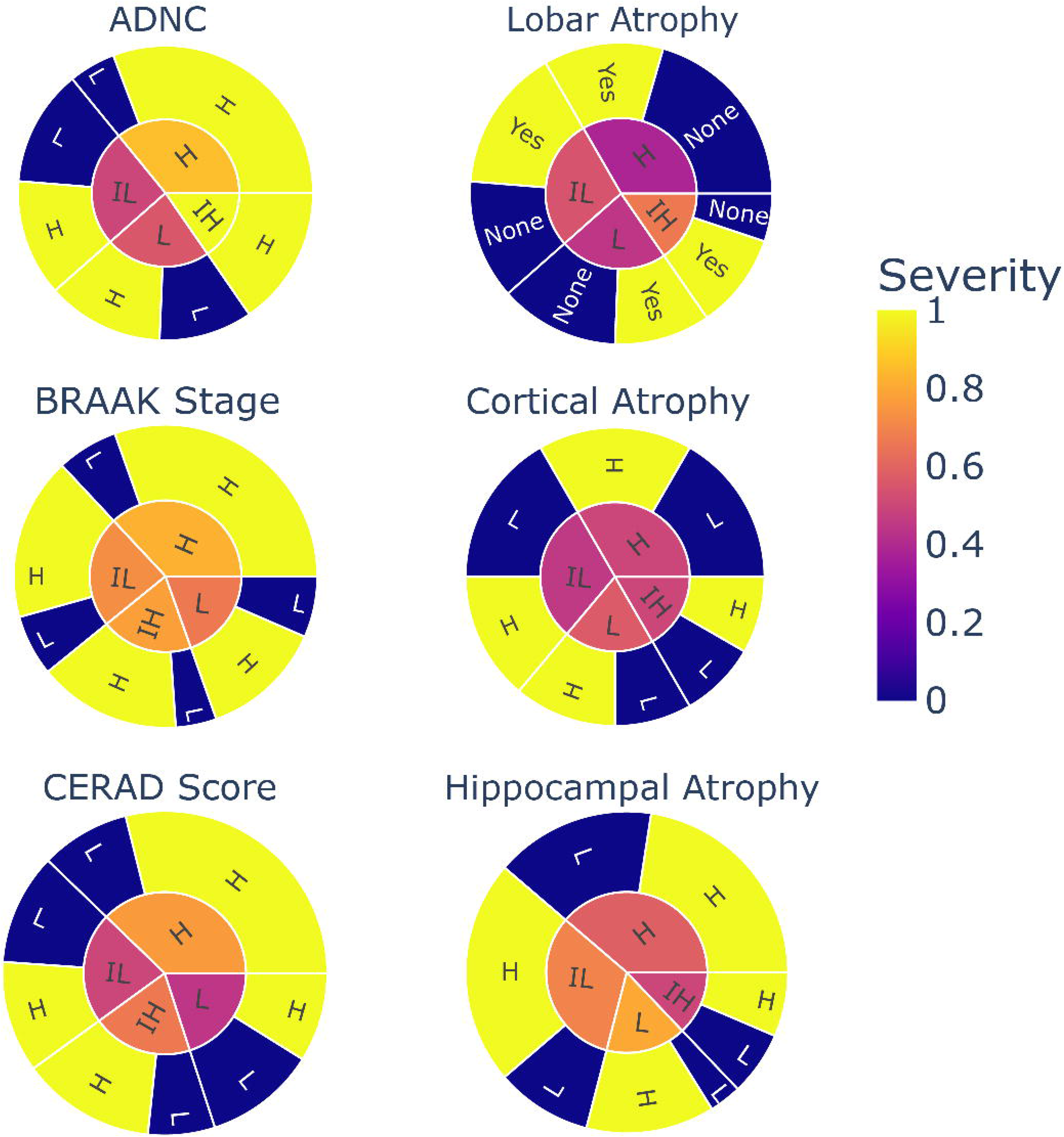
Subtype-specific associations with post-mortem data. Pathology at time of autopsy does not differ between the 4 risk-based subtypes (cortical atrophy: X^2^(3)=0.23, p=0.97; hippocampal atrophy: X^2^(3)=1.22, p=0.75; lobar atrophy: X^2^(3)=1.54, p=0.67; BRAAK stage: X^2^(3)=0.89, p=0.83; ADNC: X^2^(3)=7.21, p=0.065; CERAD score, X^2^(3)=3.39, p=0.34).

Therefore, we assessed the impact of a patient’s Subtype on the SHAP value, controlling for subject ID in addition to the region of the brain for each datapoint. A log transform was taken to stabilize the variance of the data. Brain overlays of each top atrophy-related region were plotted using the *plot_stat_map* function from the Nilearn package (https://nilearn.github.io). Finally, the *circlize* package in R was used to plot chord diagrams (Gu et al. 2014) in conjunction with the *viridis* package for colorization. To construct the chord diagrams, we obtained the union of the regions with the 5 top values of atrophy-related importance for each of the subtypes, totalling 11 regions. We determined the partial correlation coefficients between SHAP values for each of these 11 regions for each subtype and plotted the 3 regions with the highest partial correlation coefficients, and 3 regions with the lowest partial correlation coefficients. The *centrality* of each region, as measured by the sum of the absolute values of all incident edge partial correlations, determined the width of each region. The width of each edge was determined by the magnitude of the connection’s partial correlation coefficient.

### Neuropathological analysis

For the pathology analysis, we utilized sunburst plots from the *plotly* library, and performed this analysis only for our NACC data. Due to the relatively small numbers of patients available, we consolidated pathological classifications. For *cortical atrophy*, we grouped together *moderate* and *severe* atrophy as “high”, and *mild* atrophy as “low”. A similar method was utilized for Hippocampal Atrophy. For BRAAK staging, Stages 0-2 were considered “Low”, while Stages 3 or above were considered “high” stages. For ADNC scores, “Not AD” and “Low” probability of AD were considered “lower” probability, whereas “Intermediate” and “High” probabilities were grouped as “higher” probabilities of AD. for CERAD scores, C0-C1 were considered “low” scores, whereas C2-C3 were considered “high” scores. Lobar Atrophy was only scored as Present or Absent, so these were not grouped any further. The total numbers of patients with pathology data were between 31 and 46 for each of these 4 types of pathology.

## Results

### Demographic analysis

NACC and ADNI data were heterogeneous, where subjects in the NACC database progressed to AD more rapidly than those in the ADNI dataset (p<0.005) (Fig. 1A). The more rapid progression from MCI to AD in NACC was associated with lower MMSE scores at baseline when pooled compared to ADNI (p<0.001). In both datasets, lower MMSE scores was associated with more rapid progression to AD. Interestingly, the average age in the NACC cohort was higher than that of the ADNI cohort (p=0.03), though older age generally corresponds to lower rates of AD progression.

The CSF biomarker findings from the ADNI dataset were similar to those found in many previous studies. Aβ42 levels are higher in persons who take longer to progress to AD, and t-tau and p-tau are lower in persons who take longer to progress to AD. Also of note is the large variability in these CSF values within and between datasets, which could reflect person as well as collection variability. NACC and ADNI cohorts did not differ in persons in terms of number of ApoE4 alleles, sex, and proportions of subjects progressing to AD within 2, 4, and greater than 4 years.

### Patterns of risk-based subtypes during MCI to AD conversion

Mean z-scored gray matter volumes (GMVs) of various brain regions of interest (ROIs) were calculated as described in the methods for each of the 4 subtypes: high (H), intermediate-high (IH), intermediate-low (IL), and low (L). GMVs were found to be highly correlated within subtypes over time and less correlated with different subtypes (H vs H, Spearman correlation: ρ=0.85, p<0.001; IH vs IH: ρ=0.73, p<0.001; IL vs IL, ρ=0.70, p<0.001; L vs L, ρ=0.87, p<0.001). H and IH subtypes had the greatest amount of atrophy throughout the brain in comparison to IL and L subtypes (Fig. 3A, Linear mixedeffects model, IL vs H, Z = 8.3, p<0.001; IL vs IH, Z=7.1, p<0.001; L vs IH, Z=10.8, p<0.001; H vs L, Z=12.8, p<0.001). The H subtype showed the greatest amount of atrophy in the medial temporal lobe at both initial and final timepoints (Figs. 3A & 3C). This finding correlated with persons of the H subtype having high risk of progression to AD.

We found the subtypes to be robust and consistent across time from the initial MCI diagnosis to progression to AD (Fig. 3A). Persons tended to demonstrate brain atrophy in similar patterns to remain in the same subtype. Overall, the H subtype was the most stable, with 59/60 high risk MCI subjects transitioning to high risk at time of progression to AD. Interestingly, almost all conversions of risk subtypes were to higher risk subtypes, with only 1 subject each transitioning from H to IH and from IH to IL. The L subtype was the most labile, with 9/18 transitioning to higher risk subtypes.

In subgroup L, z-scored normalized GMVs in the occipital lobe and temporal lobe (both medial and overall) were relatively large compared most other lobes (OL vs Cingulate, Basal Ganglia, Parietal Lobe, and Subcortical Area, all p < 0.05; medial temporal lobe vs Parietal Lobe, Subcortical Area, Frontal Lobe, Basal Ganglia, Cingulate, all p < 0.05 [linear mixed-effects model, p-values adjusted via Dunn-Sidak method, degrees of freedom asymptotic due to large sample size]). In subtype H, the normalized GMV of the occipital lobe was only significantly larger than that of the hippocampus (p < 0.05). Subtype IL demonstrated a larger normalized GMV in the cingulate cortex compared with the rest of the cohort (Cingulate cortex vs Temporal lobes (medial and other), subcortical regions, occipital lobe, insula, all p < 0.05; vs Frontal lobe, Parietal lobe p> 0.05), and local decreases in the occipital lobe and temporal lobe compared with the other subtypes. Subtype H demonstrated a profound decrease in normalized GMV in the medial temporal lobe relative to other lobes (comparisons vs all other lobes, p < 0.05). Of course, we must interpret these relative changes within the context of a global decrease in gray matter atrophy amongst patients with the highest risk of AD.

### Distribution of risk-based subtypes

We identified 4 subgroups of MCI patients that demonstrated distinct progression projections to AD. GMVs for subjects in each subgroup were found to be high correlated between NACC and ADNI (Fig. 3A). We calculated the risk of progression from MCI to AD for each subtype in ADNI and NACC at various timepoints after being diagnosed with MCI (Fig. 3B). In the ADNI dataset, high-risk subjects were found to have a significantly greater risk of AD progression compared with intermediate-high risk subjects at times 24mo and 48mo (X^2^(1)=7.02, p=0.008, X^2^(1)=4.84, p=0.028). While intermediate-high and intermediate-low subjects did not initially differ in progression risk at time 24mo, they were found to have different progression risk when assessed at 48mo (X^2^(1)=5.59, p=0.018). Intermediate-low and low risk subjects were found to have significantly different progression risk profiles at both 24mo (p=0.0016) and 48mo (p=0.028) after MCI diagnosis. Interestingly, the differences in progression risk seemed to decrease as time progressed to 96mo after diagnosis. Risk of progression was found to be significantly different when comparing the higher risk subtypes (high, intermediate-high) with lower risk subtypes (intermediate-low, low) at 96mo.

Similar differences in progression were found with the NACC dataset. As with ADNI subjects, high risk subjects had significantly greater probability of progression than intermediate-low (p=0.006) and low risk (p=0.002) subjects at 24mo. Curiously, high and intermediate-high risk subjects only demonstrated significant differences in risk profiles starting at time 48mo (p=0.002), while only showing a strong trend towards significance at time 24mo (p=0.08). Additional significant differences in risk were found only between high and intermediate-low and low risk subjects at 48mo. Overall, all but the highest risk subtype tended to cluster together in terms of progression risk in NACC in contrast with ADNI.

### Spatial signatures of risk-based subtypes

We extracted the top 5 brain regions with normalized GMVs most positively correlated with their respective SHAP values from our MLP model for each subtype (Fig. 5A). In other words, we selectively identified regions where the importance of a region was most dependent on its input. In subtype L, we found the region importance to be on average more positive for the top 5 regions in contrast with more negative importance in subtype H; specifically, we found SHAP values for subtype H to be significantly lower than those in subtypes IL (z=-3.008, p=0.0157) and L (z=-7.609, p<0.0001) for the regions under investigation. H and IH subtypes did not differ in terms of average SHAP values (z=-1.034, p=0.8835). SHAP values in IH were significantly smaller than in L (z=- 6.593, p<0.0001) and in IL compared to L (z=-3.848, p=0.0007), indicating that the regions with the strongest relationship to their input values in these groups indicated a below average risk for progression.

In terms of region-specific findings, GMV of the superior temporal gyrus (STG) was strongly related to the contribution of this region to AD risk across all subtypes, though only a single edge emanates from this node over all three chord diagrams, in the intermediate-high risk group (Fig. 5Cii). Here, the importance of the STG, an area of the brain that contributes to language, appears inversely related to the importance of the superior frontal gyrus, which is related to behavior (Cajanus et al. 2019). The inferior temporal gyrus and angular gyrus were in the top 5 regions for both H and IH subtypes. In addition, their importance was highly correlated as shown in Fig. 5C, and they demonstrated a direct, positive connection in the IH subtype and a connection separated by a single node, the fusiform gyrus, in the H subtype. Finally, the amygdala was an independent predictor of risk in three out of the four subtypes, and had the highest centrality in subtypes L and IL, the 4^th^ highest in subtype IH, and 2^nd^ highest in subtype H. Therefore, not only is amygdala GMV an important positive predictor of survival itself in most subtypes, but its relationship with other important regions is a feature shared by these subtypes. On average, subtypes shared regions with a similarity of 0.25±0.05 (Jaccard similarity, mean ± standard deviation).

### Deep learning models

We trained multiple deep learning probabilistic risk prediction models to predict progression of MCI to AD using both ADNI (internal dataset) and NACC data (external dataset). We compared the performance of these models based on concordance indices (CI) and Brier scores (BS). Higher CI and lower BS correlate with increased prediction accuracy of probabilistic risk prediction models. The highest performing model on our external dataset was the SCNN model utilizing transfer learning and some fixed layers, with a CI of 0.681 and BS of 0.095 (Table 1). While the base SCNN seemed to have a comparable CI to the SCNN with transfer learning and 2 unfrozen layers when evaluated on our internal dataset, transfer learning helped to increase the CI of the SCNN model between 10-19% and to decrease the BS between 24-34% on the external dataset. We saw a similar pattern with our traditional CNN models, where transfer learning helped increase the progression prediction accuracy from 0.72 to 0.98.

**Table 1.**

|  | Layers | Loss | Accuracy | CI_ADNI | CI_NACC | BS_ADNI | BS_NACC | Note |
| --- | --- | --- | --- | --- | --- | --- | --- | --- |
| SCNN | 2 | Surv | N/A | 0.742 ± 0.058 | 0.572 ± 0.039 | 0.228 ± 0.036 | 0.143 ± 0.028 | |
| SCNN_T | ~ | ~ | ~ | 0.765 ± 0.068 | 0.631 ± 0.038 | 0.259 ± 0.037 | 0.108 ± 0.013 | Transfer learning |
| SCNN_T_F | ~ | ~ | ~ | 0.729 ± 0.067 | **0.681 ± 0.002** | 0.177 ± 0.023 | **0.095 ± 0.005** | last layer unfrozen; ~25% faster |
| SCNN_T_F | ~ | ~ | ~ | 0.766 ± 0.078 | 0.638 ± 0.025 | 0.358 ± 0.010 | 0.104 ± 0.002 | last 2 layers unfrozen |
| CNN | 3 | CE | 0.72 | N/A | N/A | N/A | N/A |  |
| CNN_P | 5 | ~ | 0.98 | ~ | ~ | ~ | ~ |  |
| SCNN_5 | ~ | Surv | N/A | 0.787 ± 0.020 | 0.654 ± 0.016 | 0.187 ± 0.038 | 0.142 ± 0.013 | |
| SCNN_5_T | ~ | ~ | ~ | 0.784 ± 0.027 | 0.642 ± 0.015 | 0.186 ± 0.070 | 0.137 ± 0.013 | |
| SCNN_Res | ~ | ~ | ~ | 0.603 ± 0.016 | 0.505 ± 0.003 | 0.232 ± 0.015 | 0.149 ± 0.014 | |
| VIT | N/A | ~ | 0.72 | N/A | N/A | N/A | N/A |  |
| SViT_Conv | ~ | ~ | N/A | 0.693 ± 0.042 | 0.570 ± 0.021 | 0.301 ± 0.043 | 0.166 ± 0.012 | Best of Conv |
| SViT_Linear | ~ | ~ | ~ | 0.697 ± 0.049 | 0.579 ± 0.049 | 0.291 ± 0.013 | 0.166 ± 0.051 | Best of Linear |
| MLP_PV | ~ | ~ | ~ | **0.869 ± 0.025** | 0.673 ± 0.008 | 0.076 ± 0.020 | 0.126 ± 0.007 | |
| MLP_C | ~ | ~ | ~ | 0.764 ± 0.059 | N/A | 0.169 ± 0.049 | N/A | |
| MLP_PVA | ~ | ~ | ~ | 0.854 ± 0.030 | ~ | **0.071 ± 0.018** | ~ | |
| MLP_PVM | ~ | ~ | ~ | 0.866 ± 0.030 | ~ | 0.072 ± 0.017 | ~ | |
| MLP_PVAM | ~ | ~ | ~ | 0.858 ± 0.035 | ~ | 0.068 ± 0.018 | ~ | |

Models incorporating the vision transformers did not perform well compared to the other tested models. These models had the lowest concordance indices and highest Brier indices when tested on both the internal and external datasets. In terms of performance on the internal dataset, the MLP models were among the highest performers. In fact, the MLP model utilizing GMVs resulted in the highest concordance index on the internal dataset of 0.87 while the MLP model using normalized GMVs, age, and MMSE scores resulted in the lowest BS of 0.068. The greatest benefit in decreasing the BS was observed when we added either age or MMSE to the MLP. Allowing the MLP to use both age and MMSE together did not seem to make as much of a difference in performance and it decreased the CI compared to the MLP using only MMSE.

### Pathological confirmation of results

Interestingly, of the patients who we were able to obtain pathology for in our validation cohort (NACC), no measures of AD-related pathology differed between subtypes, and the proportions of patients who progressed to AD versus those who remained MCI were similar in patients with pathology available (Chi-square tests for independence, all p > 0.05). We used ADNC as an aggregate measurement of Alzheimer’s Disease neuropathology (it is a composite of characteristic amyloid plaques (APs), neurofibrillary tangles (NTs), and neuritic plaques (NPs), the latter two of which translate to BRAAK staging (NFTs), CERAD score (NPs)) (Montine et al. 2012). Only 8/39 patients with data for ADNC demonstrated no AD, and of these only 25% were diagnosed as clinically having AD.

## Discussion

We demonstrated that (1) patterns of gray matter volume in risk-based subtypes are stable over time, (2) gray matter volume in pre-established risk-based subtypes correlates with similar patterns of GM volume loss in an external dataset, (3) both MLPs and sCNNs, and to a lesser extent sViTs, that are trained on GM volumes, are able to recapitulate the risk determined empirically in these datasets (ADNI and NACC), and (4) risk-based subtypes demonstrate distinct patterns of region significance, as defined by a strong positive correlation between SHAP value and gray matter volume.

Although choosing Aβ-42 as a marker for progression risk was theoretically justified (Buchhave 2012), it was unclear the extent to which different brain regions would distinguish themselves in different risk-based subtypes, and it was also unclear whether or not we would be able to capture distinct patterns of GM volume loss with this methodology or whether or not we would be capturing different stages of the same underlying process. Persons in the ADNI dataset with a screening-visit diagnosis of MCI were specifically selected to have symptoms of amnesia. As up to this point anatomical subtyping has demonstrated great success in differentiating patients with different areas of cognitive impairment (Murray et al. 2011; Varol et al. 2017), it was a strong possibility that by limiting our initial dataset primarily to patients with shared deficits in this cognitive domain, we would only isolate a single anatomical subtype at different points in time instead of multiple anatomical subtypes. We demonstrate that at least partially, these risk-based subtypes are distinct anatomically in terms of relative region size as they remain largely stable over time. While overall, persons in the higher risk groups demonstrate more global atrophy than those in the lower risk groups, each subtype has specific regions and lobes that have higher or lower volumes of gray matter when normalized to intracranial volume.

Mapping risk subtypes to the validation dataset by clustering based on GM volume centroids recapitulated broadly the survival curves in our subtyping scheme, with the highest risk patients progressing more rapidly to AD. Furthermore, our models were able to capture the survival curves of patients in both with good overall discrimination and calibration. Interestingly, our MLP model that utilized only normalized GM volumes from regions defined from an atlas significantly outperformed all SCNNs and SViTs in terms of Concordance Index and Brier Score on the training dataset (ADNI). However, on validation data, SCNNs that used transfer learning and fixed layers shared comparable CIs and Brier Scores that were superior to that of the MLP. Given that these models generalized comparably to the MLP and require less human input i.e., prior specification of brain regions, SCNNs could be a powerful tool to forecast AD progression and classify persons into higher or lower risk groups.

In order to infer the contributions of different brain regions to predicting the risk of progression to AD, we performed a SHAP analysis on our MLP model, highlighting regions whose importance was most dependent on the underlying GM volume, a measure we denote as *atrophy-related importance.* This is an intuitive measure capturing the idea that brain regions with larger underlying *normalized* gray matter volumes are more important for model predictions. If we know that the hippocampus is very large for a patient that is otherwise high-risk, this might more strongly inform the model than knowledge about a lobe of the cerebellum. Regions classically associated with AD showed strong atrophy-related importance across most subtypes, such as the amygdala (3/4 subtypes) (Poulin et al. 2011). Previous studies have highlighted the importance of amygdaloid degeneration. Mizuno and others found that atrophy of the amygdala correlated with overall memory performance in a dose-dependent manner (Jack et al. 1997; Mizuno et al. 2000). Our finding of the amygdala having high atrophy-related importance strongly supports these previous findings that the amygdala plays a crucial role in the early stages of AD development. The superior temporal gyrus, a center associated with language which has been shown to demonstrate atrophy and decreased activity in patients with AD (Harasty et al. 1999; Peters et al. 2009). Our finding of the importance of atrophy in the angular gyrus in progression to AD in the higher risk subtypes is also well known in the literature (Hirao et al. 2005; Karas et al. 2008). In particular, we found atrophy in the inferior temporal gyrus (ITG) to also be associated with higher risk of progression to AD. Synaptic loss in the ITG has been shown to occur in the earliest stages of AD progression, with amnestic MCI patients having up to 36% fewer synapses in this region compared to normal controls and lower verbal fluency scores on neuropsychological testing (Scheff et al. 2011). PET imaging has also revealed strong associations between increased FTP signal in the ITG and accelerated rates of cortical thinning (Scott et al. 2020). Our observations of atrophy in the ITG increasing risk of AD progression are consistent with these previous findings.

Pathological findings confirmed that we were, indeed, successful in subtyping and modeling patients with confirmed AD. AD pathology is evident prior to the onset of AD symptoms, and so we were even able to confirm AD pathology in our heterogeneous cohort of AD progressors and non-progressors, with no differences in patients with AD pathology between different subtypes.

Risk-based subtyping could be critical to paving a new way forward for physicians and pharmaceutical companies to provide targeted therapy for patients with mild cognitive impairment who would benefit from early intervention. The most well-established method of establishing risk of progression to AD is via measurement of CSF biomarkers such as Aβ, measurements have substantial between-lab variability. For example, even centers in the highly curated NACC dataset do not determine CSF values in a uniform fashion. In addition, obtaining CSF data is invasive and while in general lumbar punctures are low-risk procedures, they can lead to bleeding and infection. The strength of our method here is that we utilize MRI, a widely available, non-invasive investigation technique, which can risk-stratify people based on a highly standardized set of CSF data, map risk groups to imaging, and utilizing deep-learning methodologies, map risk to external datasets (NACC). Other groups have subtyped patients based on maximizing anatomical differences between brains; here, we maximize differences in risk and use anatomical differences to propagate this risk stratification to external datasets. Therefore, our method is a simple, extensible methodology that is best utilized to forecast differences in risk between different patients with AD.

## Data Availability

All data produced are available online at http://adni.loni.usc.edu and https://naccdata.org.

## Supplementary figures

**Figure S1.**
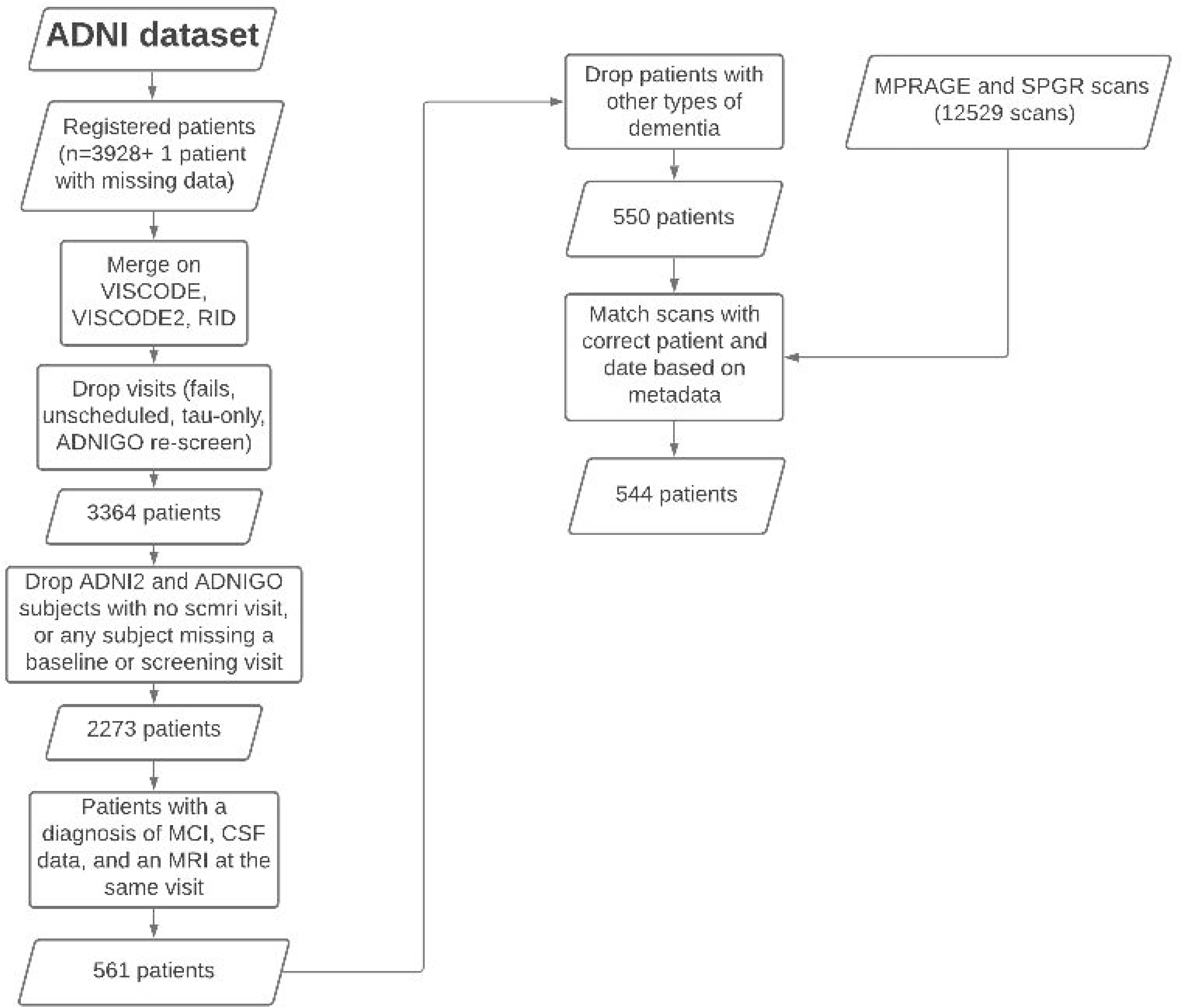
A flow chart demonstrating the process of collecting and collating metadata from the ADNI cohort and aligning the metadata with images.

**Figure S2.**
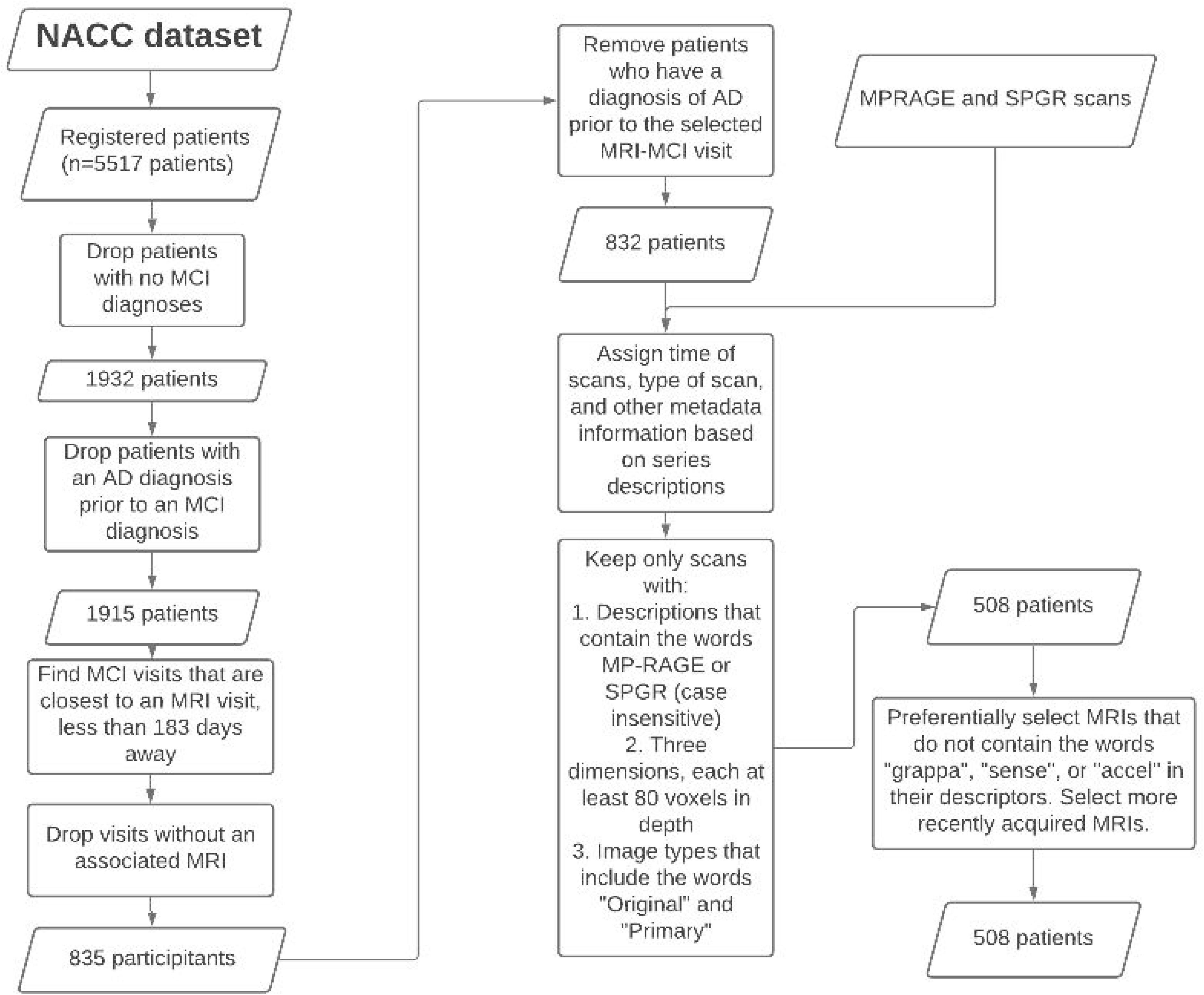
A flow chart demonstrating the process of collecting and collating metadata from the NACC cohort and aligning the metadata with images.

**Figure S3.**
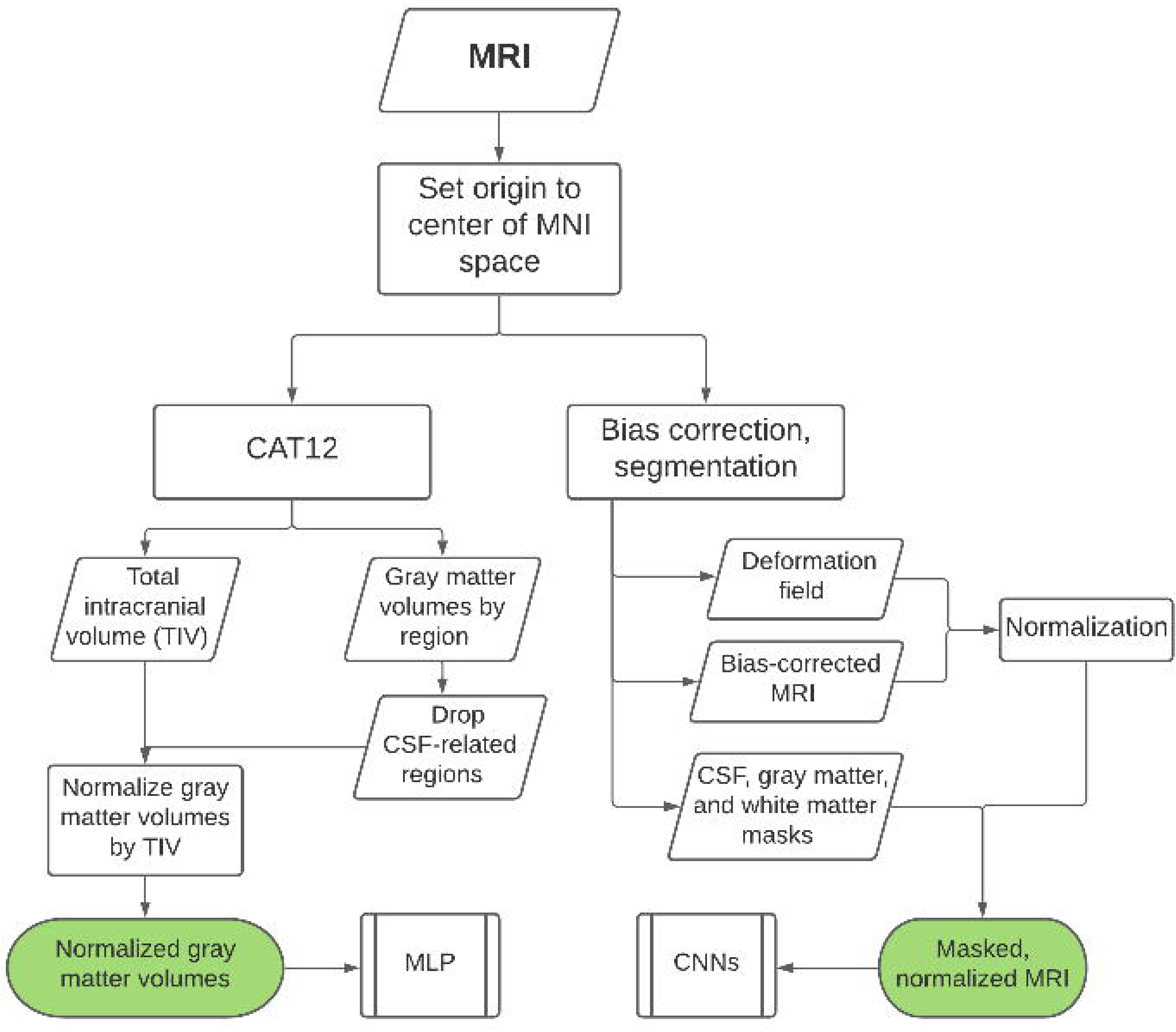
A flow chart demonstrating the two different image-processing pipelines. On the right side, we utilized a pure SPM12 pipeline in order to bias-correct and skull-strip brains to be used by the S-ViT model and S-CNN models. On the left, we obtained gray matter volumes corresponding to each region of the Neuromorphometrics atlas utilizing CAT12.7.

**Figure S4.**
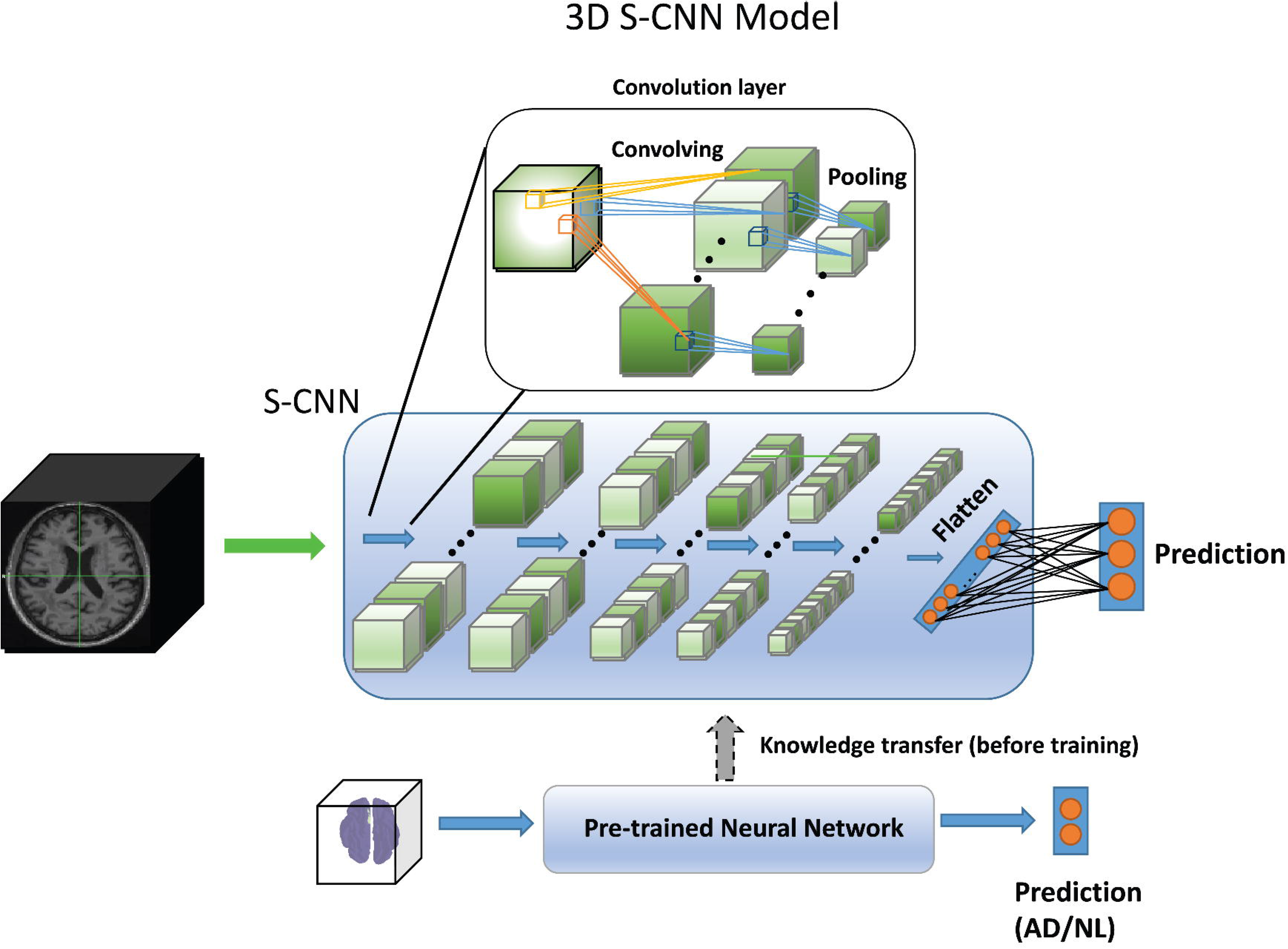
Figure of the survival convolutional neural network (S-CNN). Each convolutional layer (upper part) is composed by convolving and pooling operations, as well as dropouts. After 5 convolutional layers (middle part), the outputs were flattened and passed through a dense layer for final prediction. Before training, the network’s parameters will be initialized using a pre-trained CNN’s weights for knowledge transfer (lower part), which is trained using a different set of inputs and labels. The loss of the S-CNN is replaced by survival loss mentioned earlier.

**Figure S5.**
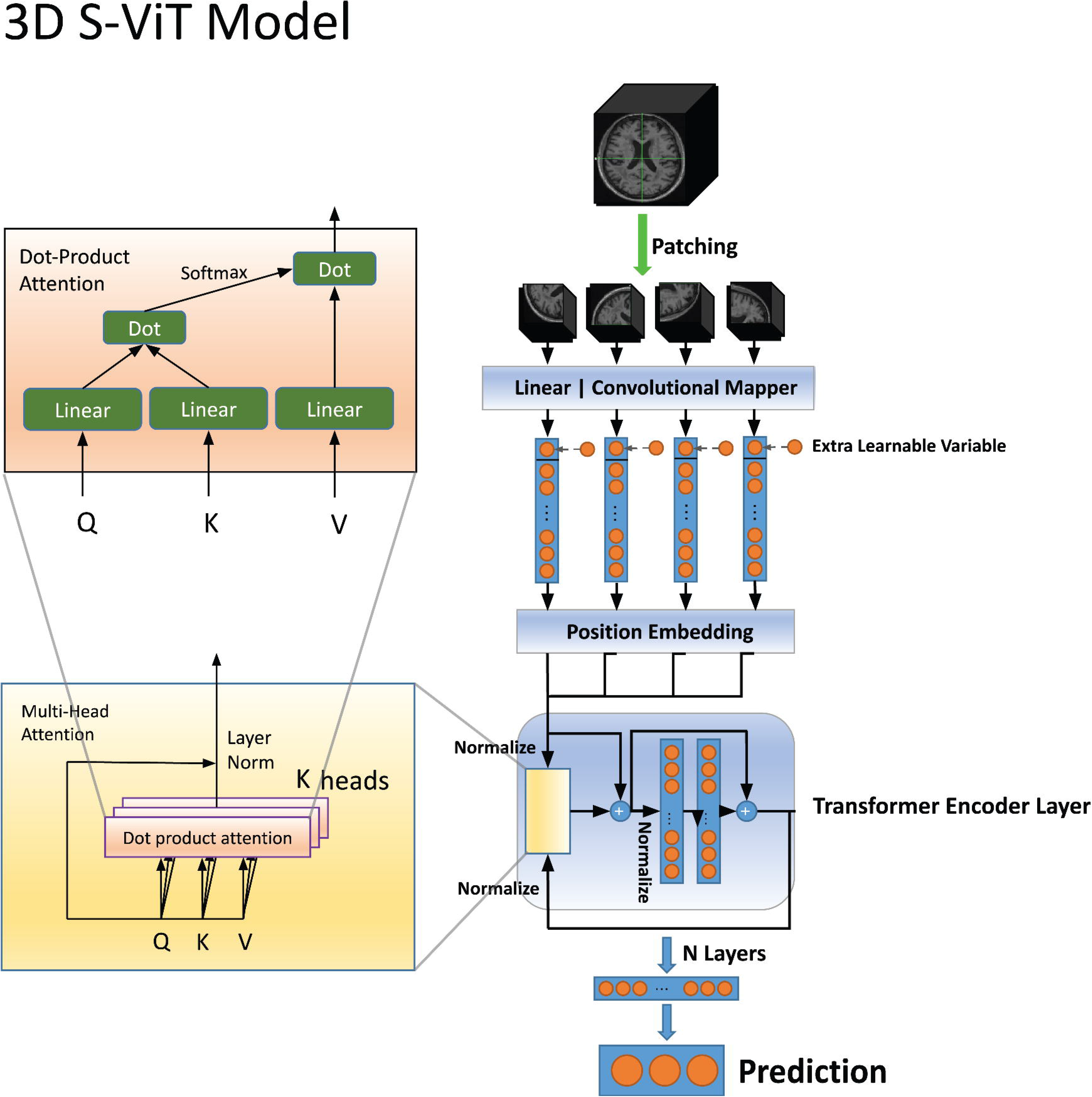
Figure of the survival vision transformer (S-ViT). The upper left figure is the self-attention, where multiple is combined concurrently in the lower left part (multi-head self-attention). The input was first divided into separate non-overlapping patches and passed through a mapper, and the resulting vectors were appended with an extra learnable variable before being sent into the positional embedder. The resulting vectors were sent into the multi-head self-attention mentioned earlier, followed by a multi-layer perceptron. After several iterations, outputs were sent into a dense layer for final prediction. Like the S-CNN, the loss of the S-ViT was also replaced by the survival loss.

**Figure S6.**
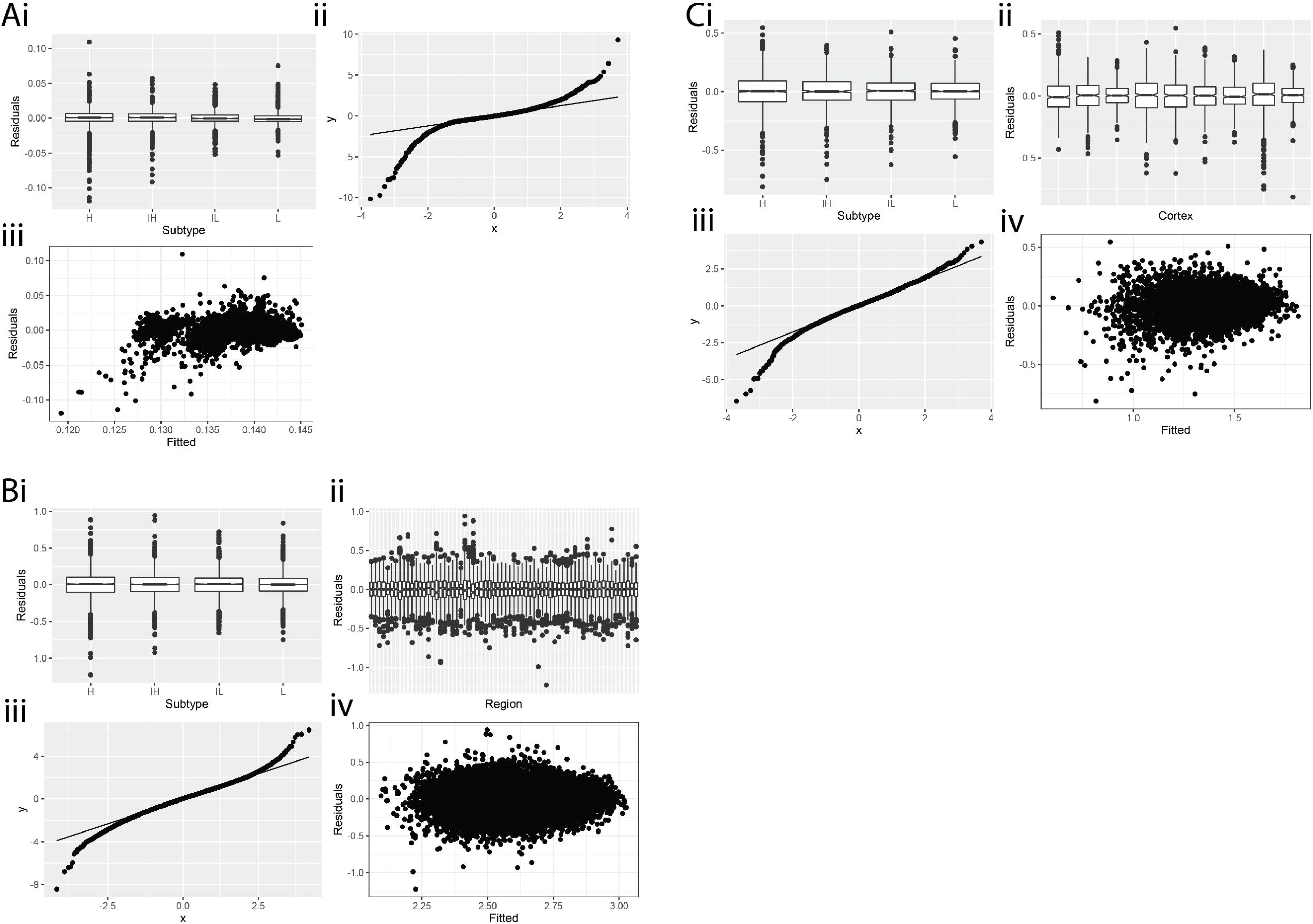
A figure demonstrating model diagnostics for the linear mixed-effects models utilized in this study. **A.** Variance in the residuals between different subtypes (i) is similar across all 4 subtypes. In the Q-Q plot (ii), there is divergence of the distribution of the residuals from a normal distribution at the tails. In (iii), it appears that the values of the residuals are mostly independent of fitted values. Subplot **B.** corresponds to Equation 1 and demonstrates similar findings, here showing consistent variance between different regions (i) and subtypes (ii). Subplot **C.** is an analogous plot that corresponds to Equation 2.

**Table S1.**
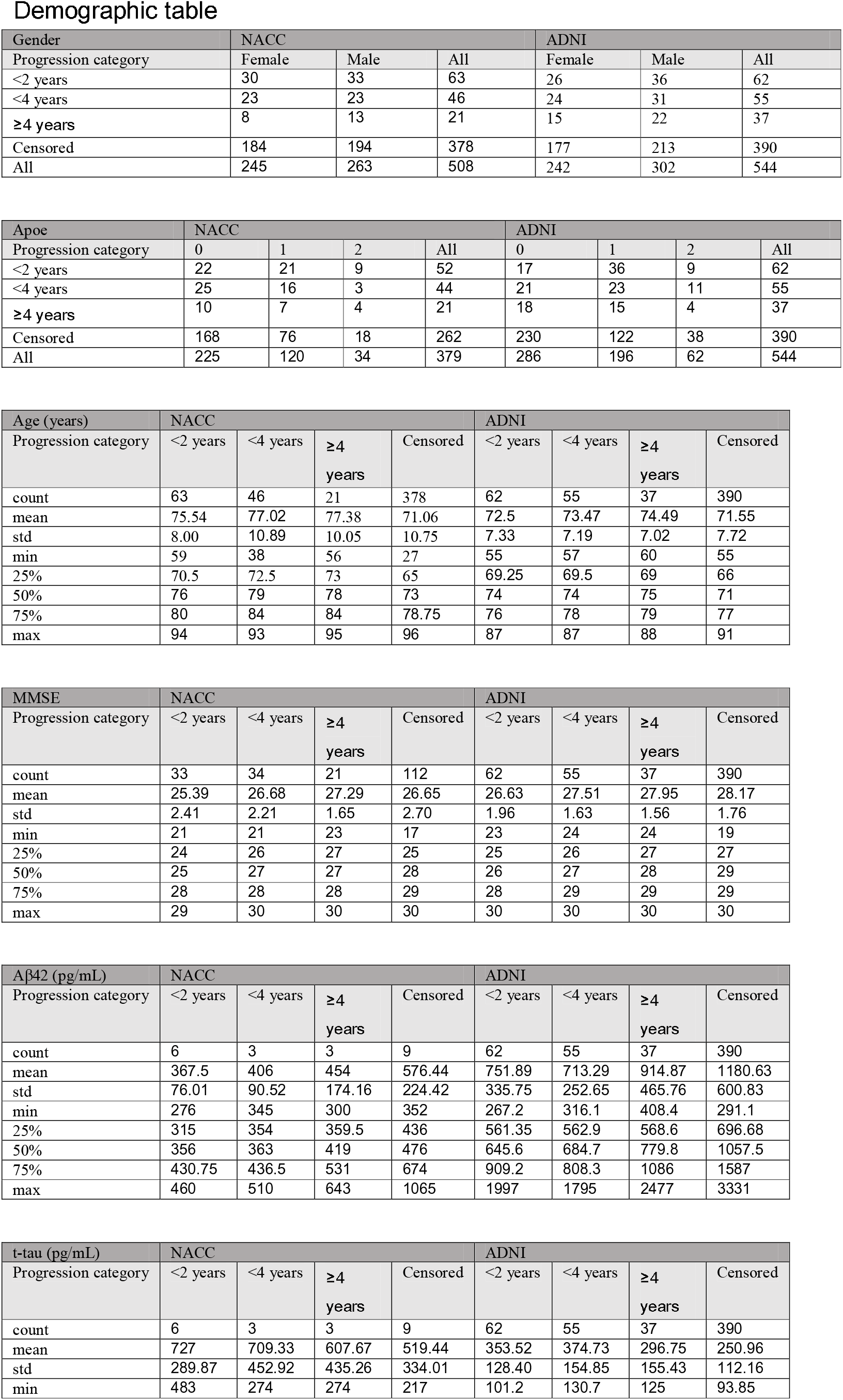
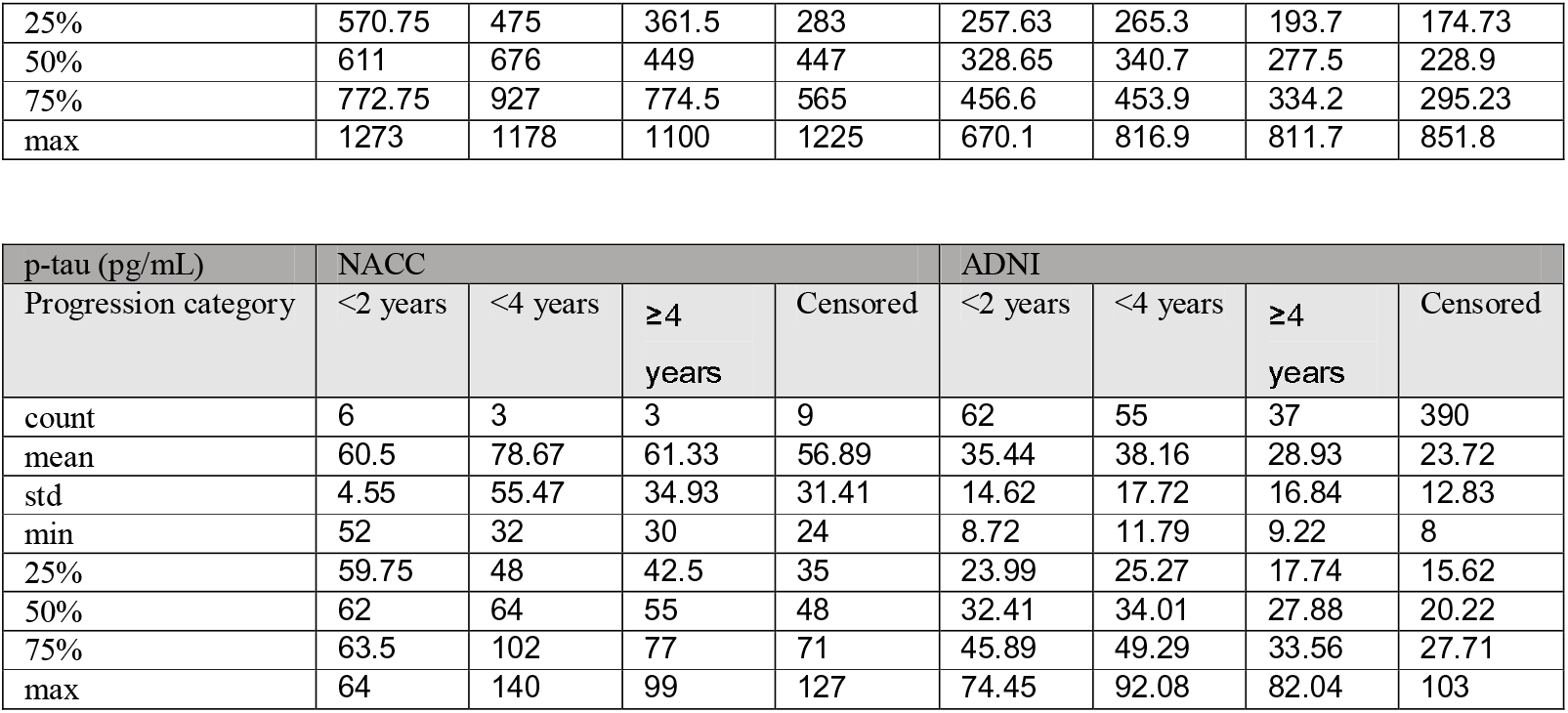
Summary statistics for both the NACC and ADNI cohorts that correspond to the figures in Figure 1. In summary, t-tau and p-tau tended to be larger in groups where patients progressed more rapidly to AD, while Aβ tended to be smaller in these groups. There were no differences between the number of APOE alleles in the average ADNI versus NACC person, and genders were also equally distributed between the two cohorts. NACC patients were in general older and had lower MMSE scores.

Demographic table
| Gender | NACC |  |  | ADNI |  |  |
| --- | --- | --- | --- | --- | --- | --- |
| Progression category | Female | Male | All | Female | Male | All |
| <2 years | 30 | 33 | 63 | 26 | 36 | 62 |
| <4 years | 23 | 23 | 46 | 24 | 31 | 55 |
| ≥4 years | 8 | 13 | 21 | 15 | 22 | 37 |
| Censored | 184 | 194 | 378 | 177 | 213 | 390 |
| All | 245 | 263 | 508 | 242 | 302 | 544 |

| Apoe | NACC |  |  |  | ADNI |  |  |  |
| --- | --- | --- | --- | --- | --- | --- | --- | --- |
| Progression category | 0 | 1 | 2 | All | 0 | 1 | 2 | All |
| <2 years | 22 | 21 | 9 | 52 | 17 | 36 | 9 | 62 |
| <4 years | 25 | 16 | 3 | 44 | 21 | 23 | 11 | 55 |
| ≥4 years | 10 | 7 | 4 | 21 | 18 | 15 | 4 | 37 |
| Censored | 168 | 76 | 18 | 262 | 230 | 122 | 38 | 390 |
| All | 225 | 120 | 34 | 379 | 286 | 196 | 62 | 544 |

| Age (years) | NACC |  |  |  | ADNI |  |  |  |
| --- | --- | --- | --- | --- | --- | --- | --- | --- |
| Progression category | <2 years | <4 years | ≥4 years | Censored | <2 years | <4 years | ≥4 years | Censored |
| count | 63 | 46 | 21 | 378 | 62 | 55 | 37 | 390 |
| mean | 75.54 | 77.02 | 77.38 | 71.06 | 72.5 | 73.47 | 74.49 | 71.55 |
| std | 8.00 | 10.89 | 10.05 | 10.75 | 7.33 | 7.19 | 7.02 | 7.72 |
| min | 59 | 38 | 56 | 27 | 55 | 57 | 60 | 55 |
| 25% | 70.5 | 72.5 | 73 | 65 | 69.25 | 69.5 | 69 | 66 |
| 50% | 76 | 79 | 78 | 73 | 74 | 74 | 75 | 71 |
| 75% | 80 | 84 | 84 | 78.75 | 76 | 78 | 79 | 77 |
| max | 94 | 93 | 95 | 96 | 87 | 87 | 88 | 91 |

| MMSE | NACC |  |  |  | ADNI |  |  |  |
| --- | --- | --- | --- | --- | --- | --- | --- | --- |
| Progression category | <2 years | <4 years | ≥4 years | Censored | <2 years | <4 years | ≥4 years | Censored |
| count | 33 | 34 | 21 | 112 | 62 | 55 | 37 | 390 |
| mean | 25.39 | 26.68 | 27.29 | 26.65 | 26.63 | 27.51 | 27.95 | 28.17 |
| std | 2.41 | 2.21 | 1.65 | 2.70 | 1.96 | 1.63 | 1.56 | 1.76 |
| min | 21 | 21 | 23 | 17 | 23 | 24 | 24 | 19 |
| 25% | 24 | 26 | 27 | 25 | 25 | 26 | 27 | 27 |
| 50% | 25 | 27 | 27 | 28 | 26 | 27 | 28 | 29 |
| 75% | 28 | 28 | 28 | 29 | 28 | 29 | 29 | 29 |
| max | 29 | 30 | 30 | 30 | 30 | 30 | 30 | 30 |

| Aβ42 (pg/mL) | NACC |  |  |  | ADNI |  |  |  |
| --- | --- | --- | --- | --- | --- | --- | --- | --- |
| Progression category | <2 years | <4 years | ≥4 years | Censored | <2 years | <4 years | ≥4 years | Censored |
| count | 6 | 3 | 3 | 9 | 62 | 55 | 37 | 390 |
| mean | 367.5 | 406 | 454 | 576.44 | 751.89 | 713.29 | 914.87 | 1180.63 |
| std | 76.01 | 90.52 | 174.16 | 224.42 | 335.75 | 252.65 | 465.76 | 600.83 |
| min | 276 | 345 | 300 | 352 | 267.2 | 316.1 | 408.4 | 291.1 |
| 25% | 315 | 354 | 359.5 | 436 | 561.35 | 562.9 | 568.6 | 696.68 |
| 50% | 356 | 363 | 419 | 476 | 645.6 | 684.7 | 779.8 | 1057.5 |
| 75% | 430.75 | 436.5 | 531 | 674 | 909.2 | 808.3 | 1086 | 1587 |
| max | 460 | 510 | 643 | 1065 | 1997 | 1795 | 2477 | 3331 |

| t-tau (pg/mL) | NACC |  |  |  | ADNI |  |  |  |
| --- | --- | --- | --- | --- | --- | --- | --- | --- |
| Progression category | <2 years | <4 years | ≥4 years | Censored | <2 years | <4 years | ≥4 years | Censored |
| count | 6 | 3 | 3 | 9 | 62 | 55 | 37 | 390 |
| mean | 727 | 709.33 | 607.67 | 519.44 | 353.52 | 374.73 | 296.75 | 250.96 |
| std | 289.87 | 452.92 | 435.26 | 334.01 | 128.40 | 154.85 | 155.43 | 112.16 |
| min | 483 | 274 | 274 | 217 | 101.2 | 130.7 | 125 | 93.85 |

|  |  |  |  |  |  |  |  |  |
| --- | --- | --- | --- | --- | --- | --- | --- | --- |
| 25% | 570.75 | 475 | 361.5 | 283 | 257.63 | 265.3 | 193.7 | 174.73 |
| 50% | 611 | 676 | 449 | 447 | 328.65 | 340.7 | 277.5 | 228.9 |
| 75% | 772.75 | 927 | 774.5 | 565 | 456.6 | 453.9 | 334.2 | 295.23 |
| max | 1273 | 1178 | 1100 | 1225 | 670.1 | 816.9 | 811.7 | 851.8 |

| p-tau (pg/mL) | NACC |  |  |  | ADNI |  |  |  |
| --- | --- | --- | --- | --- | --- | --- | --- | --- |
| Progression category | <2 years | <4 years | ≥4 years | Censored | <2 years | <4 years | ≥4 years | Censored |
| count | 6 | 3 | 3 | 9 | 62 | 55 | 37 | 390 |
| mean | 60.5 | 78.67 | 61.33 | 56.89 | 35.44 | 38.16 | 28.93 | 23.72 |
| std | 4.55 | 55.47 | 34.93 | 31.41 | 14.62 | 17.72 | 16.84 | 12.83 |
| min | 52 | 32 | 30 | 24 | 8.72 | 11.79 | 9.22 | 8 |
| 25% | 59.75 | 48 | 42.5 | 35 | 23.99 | 25.27 | 17.74 | 15.62 |
| 50% | 62 | 64 | 55 | 48 | 32.41 | 34.01 | 27.88 | 20.22 |
| 75% | 63.5 | 102 | 77 | 71 | 45.89 | 49.29 | 33.56 | 27.71 |
| max | 64 | 140 | 99 | 127 | 74.45 | 92.08 | 82.04 | 103 |

## Notes

### Competing Interest Statement

The authors have declared no competing interest.

### Funding Statement

This project was supported by grants from the Karen Toffler Charitable Trust, the Michael J Fox Foundation, the Lewy Body Dementia Association, the Alzheimers Drug Discovery Foundation, the American Heart Association (20SFRN35460031), the National Institutes of Health (R21-CA253498, RF1-AG062109, RF1-AG072654, U19-AG065156, P30-AG066515, R01-NS115114, K23-NS075097, U19-AG068753 and P30-AG013846), and the Hariri Institute for Computing and Computational Science & Engineering at Boston University.

### Author Declarations

This study involves only openly available human data, which can be obtained from the Alzheimers Disease Neuroimaging Initiative and the National Alzheimers Coordinating Center. Links: http://adni.loni.usc.edu and https://naccdata.org.

