## Supplement for "Deep learning-driven risk-based subtyping of cognitively impaired individuals"

**Supplementary material**

In this file, we provided additional details related to our methods, including the multi-layer perceptron (MLP), survival convolutional neural network (S-CNN), and survival vision transformer (S-ViT).

*Transfer learning*:

Transfer learning is a method that initializes model A’s weights using a pre-trained model B’s weights instead of random initialization. This way model A starts training from the point B achieved. This method has been proven useful and is widely used in many tasks, as well as in our models. In our S-CNN model, transfer learning improves the performance of by about 2 percent in the validation set, as can be seen in table 1 provided.

*Multi-head self-attention*:

The Survival Vision Transformer (S-ViT) is a deep learning model developed based on (Alexey Dosovitskiy, 2021)’s work, where they introduced Vision Transformer for 2D image-related task. In our setting, since the input is 3D MRI scan, we modified the image-processing step and replaced the loss to survival loss, which will be described in later part of this document. S-ViT is composed by the multi-head self-attention and 2 dense layers, where the multi-head self-attention is defined as below:

$$MSA(z)=[SA_{1}(z);...;SA_{k}(z)]U_{msa}$$

$$SA(z)=Av$$

$$A(z)=softmax(qk^{T}/\sqrt{D_{h}})$$

$$[q,k,v]=zU_{qkv}$$

Where z is the input sequence, q for query, k for key, v for value, A the weight matrix such that $A_{ij}$ stands for the similarity between i and j (Alexey Dosovitskiy, 2021).

*Cross Entropy Loss*:

When pre-training for the survival convolutional neural network (S-CNN), we used the same structure except the final layer, which is different due to the different labels. Additionally, we used the classic cross entropy loss as the loss function for the pre-trained network. The cross-entropy loss function is defined below, where log softmax and negative log likelihood are combined together:

$$Loss_{CE}(x, class)=weight[class](-x[class]+log\left( \sum_{j} exp\left( x\left[ j \right] \right) \right)$$

the averaged loss is then

$$Loss_{CE}=\frac{\sum_{i} Loss_{CE}\left( i, class\left[ i \right] \right)}{\sum_{i} weight\left( class\left[ i \right] \right)}$$

for more details, refer to (Paszke, 2019).

*Binary Cross Entropy Loss*:

Similarly, when pre-training for the S-ViT model, we changed the final layer to align the output with the labels, and used the binary cross entropy loss defined as below:

$$Loss_{BCE}(x,y)=mean(L)=mean([l_{1},...l_{n}])$$

where,

$$l_{n}=-\omega_{n}[y_{n}*logx_{n}+(1-y_{n})*log(1-x_{n})]$$

(Paszke, 2019). However, we didn’t observe a noticeable improvement in model performance when we pre-training for S-ViT, therefore, the transfer-learning results are omitted in the manuscript.

*Survival Loss*:

All of the final deep learning models we used for predicting the MCI to AD progression applies the survival loss instead of classical loss (i.e., cross-entropy loss). The survival loss is a theoretically justified loss function, which was adopted from (Gensheimer, 2019):

$$Loss_{j}=\sum_{i}^{d_{j}} ln\left( {h_{j}}^{i} \right)+\sum_{i=d_{j}+1}^{r_{j}} ln\left( 1-{h_{j}}^{i} \right)$$

where j stands for time interval j, ${h_{j}}^{i}$ is the disease hazard probability for individual i during time interval j (provided this individual didn’t progress yet), there are $r_{j}$ individuals ‘in view’ during the interval j (i.e., didn’t progressed at beginning of j), and the first $d_{j}$ of them progressed during this interval.

The total loss is then $Loss_{S}=\sum_{j} Loss_{j}$ (Sum of loss for each time interval).

This equation incorporates time-varying baseline hazard rate and non-proportional hazards. The network that applies this loss has 3-dimensional output in our case, which corresponds to the 3 time intervals. The output is basically a separate hazard rate for each time interval.

On the other hand, according to the authors (Gensheimer, 2019):

1. It is theoretically justified and fits into the established literature on survival modeling

2. The loss function depends only on the information contained in the current mini-batch, which enables rapid training with mini-batch SGD and application to arbitrary-size datasets

3. It is flexible and can be adapted to specific situations.

It is worth noting that the time intervals are left-inclusive (i.e. [0,12), [12,24), etc.). When predicting that individual i survive at least to the end of interval j can be formulated as $S_{j}=\prod_{i=1}^{j} \left( 1-h_{i} \right)$. Clearly, to compute the likelihood of i that progressed at j, the formula is $lik=h_{j}\prod_{i=1}^{j-1} \left( 1-h_{i} \right)$. For additional details, refer to (Gensheimer, 2019).
